# Bimodal Contact Reductions and Social Homophily during COVID-19

**DOI:** 10.1101/2025.07.10.25331264

**Authors:** Sydney Paltra, Leonard Stellbrink, Jens Friedel, Mirjam E. Kretzschmar, Maged Mortaga, Kai Nagel, Hendrik Nunner, André Calero Valdez, Viola Priesemann

**Affiliations:** Institute of Land and Sea Transport Systems, Technical University Berlin, Germany; Institute of Multimedia and Interactive Systems, University of Lübeck, Germany; Max-Planck Institute for Dynamics and Self-Organization, Göttingen, Germany.; Julius Center for Health Sciences and Primary Care, University Medical Center Utrecht, Utrecht University, Utrecht, The Netherlands; Centre for Complex System Studies (CCSS), Utrecht University, Utrecht, The Netherlands; Interdisciplinary Center for the Mathematical Modeling of Infectious Disease Dynamics (IMMIDD), University of Münster, Germany; Faculty of Physics, Georg-August-University Göttingen, Germany

**Keywords:** COVID-19, Contact Patterns, Contact Reductions, Social Homophily, Heterogeneity of Contacts

## Abstract

**Background:** The COVID-19 pandemic disrupted social life and forced people to reconsider how, where, and with whom to spend time. These decisions are deeply personal and their intricacies are still poorly understood.

**Methods:** To understand how people make such decisions, we conducted an online survey in summer 2023, collecting self-reported absolute contact numbers for four time points: 2019, 03/2020, summer 2021, and 01/2023. We analyzed the resulting contact data, focusing on the quantification of heterogeneities in reductions.

**Results:** Analysis of the survey data revealed that the COVID-19 pandemic triggered substantial reductions in both the work and the leisure context. Mean reductions gradually decreased as time progressed, but by 01/2023 contact numbers remained below pre-pandemic levels. We found contact behavior to demonstrate heterogeneity in three different aspects. First, the distribution of contact reductions followed a bimodal pattern, with a distinct peak at either extreme: A large fraction of the survey participants initially strongly reduced their contacts, a smaller group maintained nearly normal contact levels, and the remainder of participants reduced their contacts intermediately. Consistent with the decrease of mean contact reductions, the relative sizes of these behavioral groups shifted over time, with participants relaxing their reductions incrementally. Second, we found risk perception to be an indicator for the strength of contact reductions: Risk-averse participants reduced their leisure contacts significantly more than risk-tolerant participants, resulting in a trend of both fewer and later COVID-19 infections. Neither age, gender, nor having a COVID-19-relevant comorbidity significantly influenced self-reported contact reductions. Third, the survey results provide evidence that social homophily persisted during the COVID-19 pandemic, revealing a correlation between participants’ and their closest contacts’ number of contacts during the COVID-19 pandemic. Risk-averse participants hereby especially preferred to maintain contact with equally careful individuals.

**Conclusions:** Our study emphasizes the time-dependency and heterogeneity of contact reductions. On the one hand, our findings can easily be integrated into epidemiological models, improving their accuracy and predictive power. On the other hand, the results may guide the design of effective public health interventions, and help to predict and understand their effectiveness.

## Introduction

Contact behavior substantially influences infectious disease spread. Whenever humans meet in person, converse, or touch, they create opportunities for pathogens to spread from one human to another. The frequency, duration, and proximity of these interactions directly affect the transmission rates [Smi09, DCZMM14]. While changes in contact behavior serve as critical determinants of disease spread, contact reductions can introduce complexity to the infection dynamics, as people continuously adjust their behaviors in response to the evolving outbreak [Fer07, WAW^+^15, SBM10, PSCVMV15, DWC^+^22, WBC^+^23]. For effective disease mitigation, it is vital to understand the complex interplay between infectious disease spread and contact behavior.

Contact behavior can be modified both through governmental nonpharmaceutical interventions (NPIs) and voluntary behavior adaptations. Studies examining contact behavior during the COVID-19 pandemic demonstrated that the combination of mandatory NPIs and voluntary contact behavior adaptations strongly reduced the number of contacts in various contexts, including work and leisure [VHV^+^21, CWG^+^20, WGC^+^23, JCB^+^24, PBN24, DWC^+^22, DZS^+^20, BMS^+^21]. Consequently, when designing NPIs, public health officials should consider the heterogeneity of the underlying social contact network. When taking heterogeneities in social network structure and personal preferences into account, public health officials can design contact reductions that remove essential connections between people, decrease the network’s connectivity, and effectively slow down disease spread [Val12].

Contact behavior, both before and during an infectious disease outbreak, demonstrates heterogeneity in three distinct aspects. First, people’s number of contacts is not uniformly distributed across the population, but displays heterogeneous patterns. Across age groups, school-aged children have the most contacts, mainly with one another, followed by contact with their parents, while the elderly have the fewest contacts [MHJ^+^08, MWW^+^21, TRB^+^21]. This heterogeneity in contact numbers influences disease spread, as people with more contacts are the central nodes in the social contact network and may serve as transmission catalysts [WLX18]. Strategically focusing contact reductions on high-contact individuals can make interventions more effective. Second, personal attributes — including age, gender, health conditions, personal preferences, trust in institutions, and perceived risk — influence the adoption of self-protective behaviors like contact reductions. Higher *risk perception* encourages contact reductions; for example, through the avoidance of crowded public events or of people who have traveled to countries with a high incidence [BM10, BAO^+^04]. Being *female* is associated with greater engagement in self-protective behaviors as well as compliance with protective measures [Lau03, BM10, JS09, GPP^+^20]. Older *age* encourages the adoption of self-protective behaviors such as vaccination and hand washing [Lau03, HC06, JS09], as does being diagnosed with a *multimorbidity* [DMFdPCV22]. Targeting individuals receptive to contact reductions may limit the cost of NPIs without compromising the effectiveness of mitigation.

Third, people have a tendency to prefer connections with others who share similar characteristics, beliefs, and backgrounds. This “birds of a feather flock together” phenomenon, known as social homophily, shapes the contact network in the context of friendships, leisure contacts, romantic partnerships, and partly also professional relationships [MSLC01, HM17, FAA12]. Social homophily may slow down disease spread, as spread over bridging ties between clusters is less likely [WS98, BS10]. In adaptive networks, social homophily may even be decisive in stopping an outbreak early, as it requires only minor disruptions in the network structure to isolate clusters, which then act as a barrier for the disease to reach further parts of the network [NBTK22]. Yet, recent experimental work suggests that such protective behavior may come at a social cost. When people prioritize infection avoidance, they may break otherwise beneficial social ties, leading to a loss of cohesion and network fragmentation [NBC^+^23]. These findings underscore the need to better understand how the structure and dynamics of social contacts influence not only the course of an outbreak but also its broader impact on social life.

Research conducted during the early stages of the COVID-19 pandemic has conclusively demonstrated significant contact reductions during the first pandemic year [DZS^+^20, LBK^+^21, MBC^+^21]. Still, a knowledge gap persists regarding which contacts are cut and which are maintained, the heterogeneity in reduction strength, and the decision-making processes that governed these reductions. Gaining a more profound understanding of the intricacies of contact behavior during the COVID-19 pandemic is essential, as it supports epidemiological modelers in representing human behavior realistically and public health officials in designing more nuanced and thus more cost-efficient NPIs for future outbreaks.

In this study, we examine three of these intricacies, reflecting the aforementioned three aspects of heterogeneity: First, we demonstrate that the aforementioned pre-pandemic heterogeneity in the number of contacts translates to heterogeneous contact reductions during the COVID-19 pandemic. While many survey participants reduced their contacts strongly, some displayed little change, and the remainder spread out between the two extremes, resulting in a bimodal distribution of reduction. This bimodal distribution of contact reductions can be observed for both work and leisure contacts. For both contexts, the group size of participants who strongly reduced decreased over the course of the pandemic, while the group sizes of intermediate reduction and little change incrementally increased. Second, we show that stronger risk perception correlates with stronger initial contact reductions as well as stronger upkeep of these reductions over time. Third, we provide evidence for the persistence of social homophily and the importance of second-order contacts during the COVID-19 pandemic.

## Methods

### Data Collection

All data was collected via an online survey in the summer of 2023. To recruit survey participants, we partnered with five German-speaking academics, whose Twitter (now X) followers ranged from 750 to 65,000 at the time of the study. The five recruiters shared the online survey on Twitter on July 19, 2023, with one recruiter additionally sharing the survey on Mastodon. To post polls on behalf of each of the five recruiters and to ensure simultaneous posting, we developed a Twitter poll bot. This bot was set up to share the polls and the survey link in a thread (multiple linked posts), allowing participants to easily access the survey. The poll bot was only used by three of the five Twitter recruiters; the remaining two recruiters posted independently. The single Mastodon recruiter shared the survey without a bot. The survey data collection ran from July 18, 2023, to August 30, 2023. We collected data in the following order: basic sociodemographic data (age, gender), number and timing of COVID-19 infections, contacts of participants, contacts of household members, contacts of closest contacts, risk perception, vaccination(s), and additional sociodemographic data (including current occupation, education). Out of 867 started surveys, 398 were completed, and participants spent on average 13 minutes filling out the survey. The survey in the original German and the anonymized survey data are available on OSF ([FNM^+^23] and [CVFN^+^25]). We discussed the representativeness of the study population in a previous publication: The majority of survey participants was between 40 and 59-years-old, individuals who have received higher education were oversampled, while one-person households were undersampled (Supplementary Figure 1). Infection numbers and 7-Day-Incidence/100,000 of the survey participants are comparable to high-effort panel studies and official reporting numbers [MNP^+^25]. Data collection was conducted in accordance with the Declaration of Helsinki and approved by the ethics committee of the University of Lübeck.

### Data Processing

All responses were anonymized during data processing: We removed personally identifiable information, such as IP addresses and free text fields. Speeders – participants who answered the survey in less than a third of the median time – were removed. Preprocessing code is publicly available on GitHub (https://github.com/hciuse/twitter-study).

### Contact Data

We defined a “contact” as any situation in which one person comes closer than two meters to another person for at least 15 minutes. This definition ignores the presence or absence of self-protective measures (mask-wearing is provided as an example in the survey). Absolute weekly contact numbers were collected differentiated by context, namely for leisure, school, and work. Contacts were collected retrospectively, with participants being asked to report their weekly number of distinct contacts for four time points: 2019, 03/2020, summer 2021, and 01/2023. For each time point, the survey provided auxiliary information on the then-current state of the COVID-19 pandemic, including the stringency of NPIs, the then-current virus strain and the availability of the vaccine (Supplementary Table 2). Weeks were chosen as the time scale because the infectious period of SARS-CoV-2 is up to one week [BMC^+^20, HZJ^+^22]. Furthermore, contact patterns vary throughout the week and our retrospective data collection method did not allow us to distinguish between weekdays and weekends [TC12, ZPSD^+^23].

Participants were asked to report their own contacts, their household members’ number of contacts, and their (non-household) closest contact’s (CC) number of contacts (see Supplementary Section Accuracy of Participants’ Reported Contact Numbers for qualitative assessment of participants’ accuracy). The collected contact data does not allow us to determine if there exists an overlap between the participant’s and the CC’s contacts: This overlap would mean that some contacts are both contacts of the CC *and* direct contacts of the participant. In the following, for reasons of simplicity, we refer to the participants’ own reported contacts as first-order contacts and the CCs’ contacts as second-order contacts.

### Classification into Subgroups

For subgroup analyses, we classified participants according to the following four characteristics:

1. *Risk Perception.* The survey included nine items on attitudes related to COVID-19 at the end of March 2020, asking participants to compare themselves to an “average person” (Table 3). Answers were collected on a scale ranging from “a lot less” to “a lot more” (see Table 4 for original German options). In the analysis, we excluded the answer options “does not apply” and “not specified”, yielding a 7-point Likert scale. We mapped the answers “a lot less”, “less”, and “slightly less” to *−*1, “just as much” to 0, and “slightly more”, “more”, “a lot more” to +1. The scale was reversed for questions about meeting close contacts despite restrictions and feeling restricted by measures. We summed up these values to calculate a risk perception score. The risk perception score may take on integer values between *−*9 and 9, but we only obtained values between *−*6 and 9 for our sample. We binned participants into two groups: risk-tolerant (risk perception score *≤* 3) and risk-averse (risk perception score *≥* 4.) We introduced the asymmetrical split of the scale to increase the size of the risk-tolerant group and maximize differences in contact behavior and number and timing of infections between groups. Some participants did not or only partially answered the risk perception survey items. Consequently, for these participants, no risk perception score could be computed and they are classified as “No Risk Perception Score Available”.
2. *Age.* Participants were asked to report their age in years. We binned participants into three age groups: 18-39, 40-59, 60+. Underage individuals were not allowed to participate in this survey due to data protection reasons.
3. *Gender.* Data on gender was collected via a multiple-choice item. Choices were “male”, “female”, “diverse”, and “I don’t want to answer”. Subanalysis by gender only included participants who reported their gender as “female” or “male” as only seven participants reported their gender as “diverse” (< 1%), making the sample too small to give statistically meaningful insights.
4. *Comorbidities.* Participants were asked if they had been diagnosed with hypertension, diabetes, cardiovascular disease, immunodeficiency, cancer, or post-COVID-19 condition. We binned participants according to whether they reported any of these comorbidities.

### Statistical Methods

#### Distribution of Change of Contacts

Participants reported absolute contact numbers for 2019, 03/2020, summer 2021, and 01/2023. For each participant, we computed the percentage change of number of contacts in 03/2020, summer 2021, and 01/2023, relative to 2019. As a percentage change from zero cannot be computed, we excluded participants who reported zero work or leisure contacts for 2019.

### Determination of Shape of Distribution

We used Bayesian modeling to quantify the shape of the distribution of change of the number of work or leisure contacts at each time point. We used Gaussian mixture models to determine if the change in the number of work and leisure contacts followed a uni-, bi-, tri-, or quatro-modal distribution. We fitted one to four (half)-normal distributions to the data using Markov chain Monte Carlo methods. We assumed the means of the normal-distributions and the shift of the half-normal distribution to be fixed, and estimated the variances and the weights of the distributions. When fitting one distribution to the data, we used a half-normal distribution shifted to *−*100% such that it only allows values *≥ −*100%. We added a normal distribution with a mean of 0% when fitting two distributions to the data. When considering three distributions, we added an additional normal distribution centered around *−*50%, and when considering four distributions, we added another normal distribution centered around +50%. Model comparison was performed using leave-one-out cross validation. Model comparison selected the model using three distributions, consisting of a narrow half-normal distribution with a lower bound of *−*100%, an equally narrow normal distribution with a mean of 0%, and a wide and flat normal distribution covering all contact reductions in between the two extremes. Due to the pointed, narrow distributions at the two extremes, we will still refer to shape of the contact reductions as “bimodal” (see Supplementary Section Bayesian Fits for details).

### Group Assignment

We used the posterior mean estimates of the selected model to estimate group membership probabilities for the three corresponding groups. Using these probabilities, we assigned participants to the groups of “strong reduction”, “intermediate reduction”, and “little change” for every time point. Few participants increased their work or leisure contacts. These participants could not be mapped to any of the aforementioned groups and were instead mapped to the group “none”.

### Kolmogorov-Smirnov tests

To test for significant differences in the distributions of changes in work and leisure contacts across the multiple time points, we applied two-sided Kolmogorov-Smirnov tests. Analogously, to test for significant differences in distributions of changes in work and leisure contacts for each time point but between subgroups, we used two-sided Kolmogorov-Smirnov tests. Finally, to test if the distributions of the number of infections as well as the ECDF of the timing of the first infection differ significantly between subgroups, we used two-sided Kolmogorov-Smirnov tests. In each instance we, reported the appropriate significance level (0.01, 0.05, 0.1).

### Confidence Intervals

We anaylzed the discrete distribution of the number of reported COVID-19 infections (0, 1, 2, 3+) using a bar chart. For each bar, we computed 95% confidence intervals. We assumed that the proportion of participants who reported *N* infections approximates the true proportion of participants who have been infected *N* times. Similarly, for the emperical cumulative distribution function (ECDF) of the timing of participants’ first infection, we computed 95% confidence intervals. Again, we assumed that the proportion of participants who having been infected by a certain date, approximate the true participant proportion of having been infected by a certain date. In either case, we assumed the true population proportion to be the mean of a binomial distribution, and as the binomial distribution is approximately normal for large enough samples, we used z-scores when computing the 95% confidence intervals.

### Missing data

We obtained 398 completed and 867 partially filled out surveys. We also used incomplete surveys for the analysis.

### Processing of POLYMOD data

We compared the survey participants’ characteristics and their reported number of 2019 work or leisure contacts to the results obtained in the POLYMOD study for Germany [MHJ^+^08]. POLYMOD data was accessed via Zenodo [MNJ^+^20].

## Data Analysis and Availability

All analysis code is publicly available on Zenodo [Pal25]. For most data analysis we used R 4.4.1, namely packages from the tidyverse, whereas to fit two to four (half)-normal distributions to the change of number of contacts, we used Python 3.11.5, more specifically PyMC 5.10.4 [WAB^+^19, APAC^+^23].

## Results

### Contact Reductions

During the COVID-19 pandemic, self-reported work and leisure contacts decreased significantly compared to the pre-pandemic contact levels of 2019. On average, work contacts were reduced by 75% in 03/2020, by 64% in summer 2021, and by 42% in 01/2023 (Fig. 1A). The Bayesian reduction model maps participants’ reductions to the three groups “strong reduction”, “intermediate reduction”, and “little change” (see Supplementary Section Bayesian Fits for model comparison). The sizes of these groups differed from one to the subsequent time point, confirming the temporal development of contact reductions (*p* < 0.01, see Supplementary Section Kolmogorov-Smirnov tests for details). In 03/2020, more than half of the participants “strongly” reduced their work contacts (55%, Table 1). This share incrementally decreased, to 42% (summer 2021), and finally to 24% (01/2023). Complementarily, the share of participants who displayed “little change” in their work contacts increased from 16% (03/2020) to 17% (summer 2021) to 31% (01/2023). Participants typically relaxed their work contact reductions incrementally, moving from “strong reduction” to “intermediate reduction” and from “intermediate reduction” to “little change” (Fig 2A). In sum, work contact reductions were relaxed over time such that by 01/2023 only 24% of participants maintained “strong” work contact reductions, 39% “intermediate reductions”, and 32% had returned to pre-pandemic levels.

**Figure 1:**
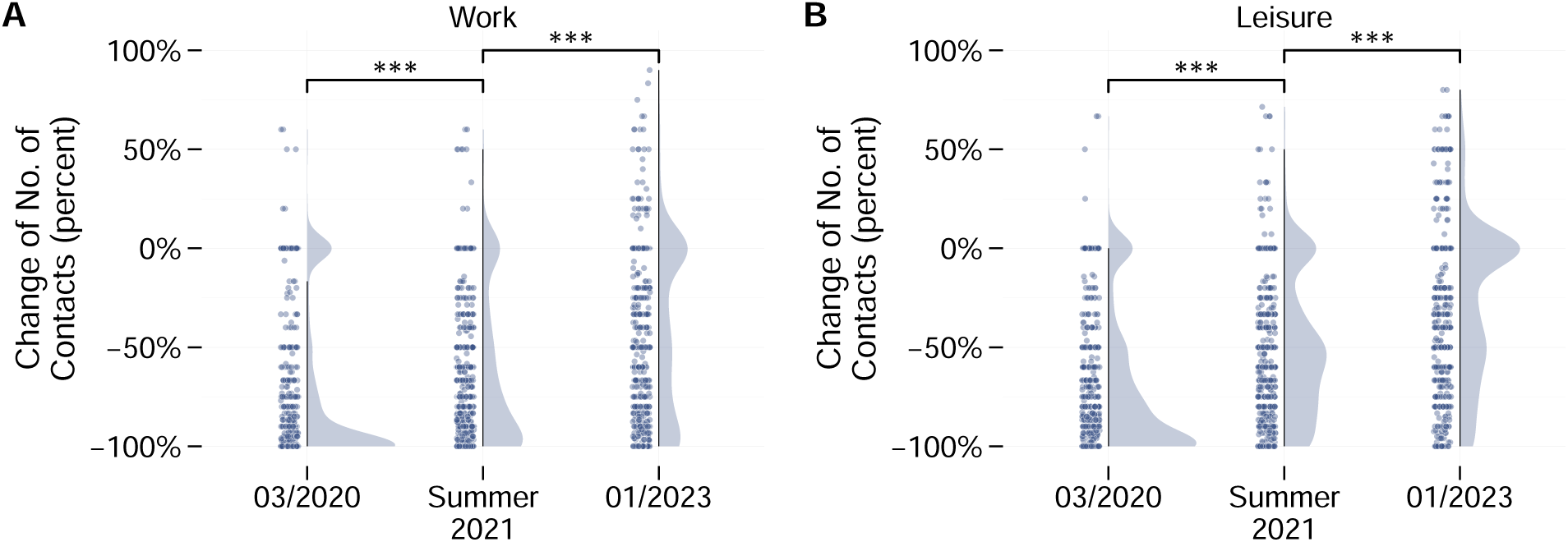
Change of number of contacts (in percent, relative to 2019). Stars indicate that the distributions differ at significance levels of \*\*\**p* < 0.01. **A. Relative number of work contacts.** On average, participants reduced their work contacts in 03/2020, summer 2021, and 01/2023. A bimodal distribution of reductions is visible for all three time points, with the group of “strong reduction” decreasing and the group of “little change” increasing as time progressed. **B. Relative number of leisure contacts.** On average, participants reduced their work contacts in 03/2020, summer 2021, and 01/2023. For all three time points, the distribution is bimodal. The share of participants who displayed little change in their leisure contacts increased over time, such that by 01/2023, around 40% of participants had returned to pre-pandemic leisure levels.

**Figure 2:**
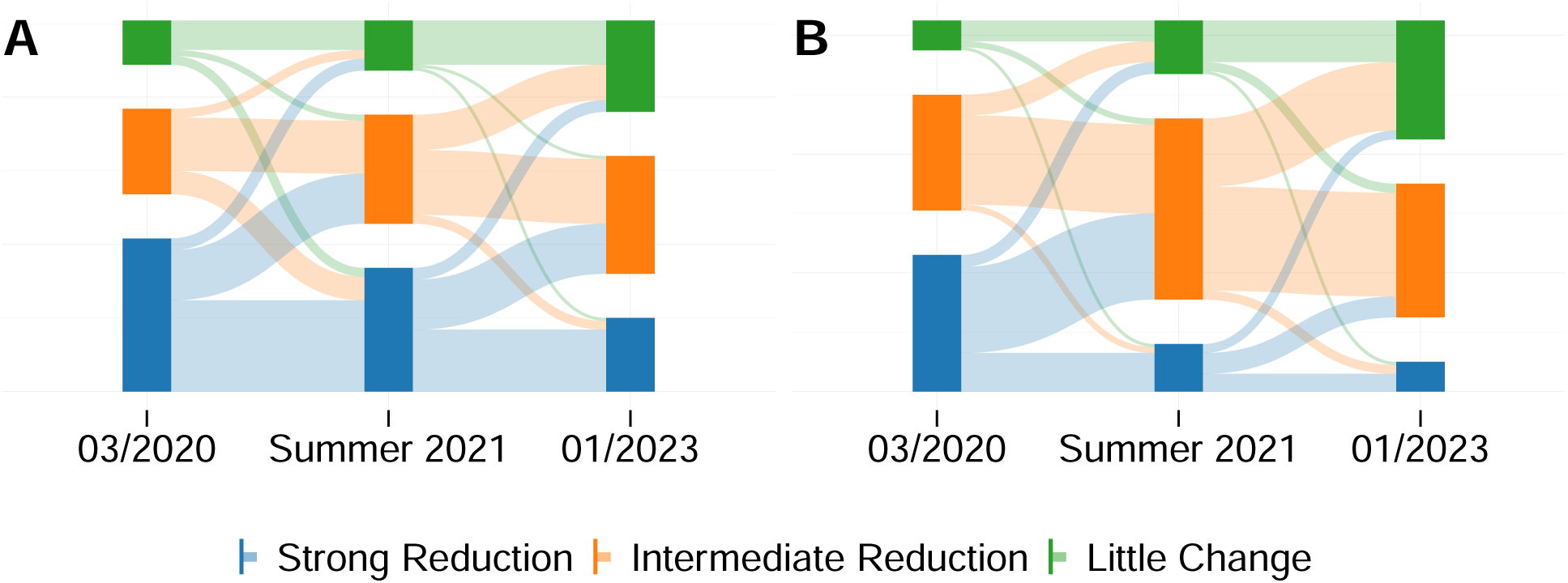
Sankey diagrams depicting the flow between reduction groups. Participants decreased their reductions over time. **Sankey diagram change of work contacts.** Participants gradually decreased their reductions (moving from “strong reduction” to “intermediate reduction” to “little change”). **Sankey diagram change of leisure contacts.** Participants gradually decrease their reductions (moving from “strong reduction” to “intermediate reduction” to “little change”). Hardly anyone moved to a more restrictive group.

**Table 1:** Group assignment based on the change of number of contacts (for details, see Subsection Statistical Methods). For both contexts, the share of individuals in the “strong reduction” group decreases over time.

|  | Work |  | Leisure |  |
| --- | --- | --- | --- | --- |
|  | N | % | N | % |
| **03/2020** |  |  |  |  |
| Strong Reduction | 401 | 55.1 | 372 | 46.5 |
| Intermediate Reduction | 203 | 27.9 | 328 | 40.9 |
| Little Change | 118 | 16.2 | 97 | 12.1 |
| None | 6 | 0.8 | 4 | 0.5 |
| **Summer 2021** |  |  |  |  |
| Strong Reduction | 301 | 42.0 | 126 | 16.1 |
| Intermediate Reduction | 281 | 39.2 | 489 | 62.6 |
| Little Change | 124 | 17.3 | 146 | 18.7 |
| None | 10 | 0.1 | 20 | 2.6 |
| **01/2023** |  |  |  |  |
| Strong Reduction | 168 | 23.6 | 60 | 8.0 |
| Intermediate Reduction | 278 | 39.0 | 340 | 45.1 |
| Little Change | 229 | 32.2 | 295 | 39.1 |
| None | 37 | 5.2 | 59 | 7.8 |

Very similar to work contacts, leisure contacts were also decreased in 03/2020, summer 2021, and 01/2023 compared to 2019. On average, participants reduced their leisure contacts by 72% in 03/2020, by 51% in summer 2021, and by 28% in 01/2023 (Fig. 1B). Analogously to work contacts, leisure contacts had not returned to pre-pandemic levels by 01/2023. The Bayesian reduction model quantifies the bimodal shape of the distribution for the three time points. The shape of the distribution changes significantly from one time point to the subsequent one (*p* < 0.01). According to the Bayesian reduction model, 47% of the participants “strongly” reduced their leisure contacts in 03/2020. The share of this group decreased over time, to 16% (summer 2021) and finally to 8% (01/2023). Again, participants incrementally relaxed their leisure contact reductions (Fig. 2B). In sum, only 8% maintained “strong” reductions, 45% intermediate reductions, and nearly 40% of participants had returned to their pre-pandemic contact patterns, highlighting a heterogeneity in contact behavior and the importance of examining contact distributions rather than just mean values.

### Subanalysis by Risk Perception Score

Risk-averse and risk-tolerant participants showed no difference in their 2019 pre-pandemic contact levels. The two groups are comparable in their general sociability, as neither the reported work nor the reported leisure contact numbers differed significantly (*p* > 0.1, Supplementary Section Pre-Pandemic Contact Data Differentiated by Risk-Perception Score).

Risk-averse participants, however, reported stronger work contact reductions than the risk-tolerant participants (Fig. 3). In 03/2020, risk-averse participants reduced their work contacts on average by 78%, while risk-tolerant participants reduced theirs by 64% (difference statistically significant, *p* < 0.05, see Supplementary Section Kolmogorov-Smirnov tests for details). Both groups relaxed their work contact reductions over time. In summer 2021, the mean work contact reduction was 65% for risk-averse and 58% for risk-tolerant participants, respectively (*p* > 0.1). In 01/2023, mean work contact reductions were 46% (risk-averse participants) and 36% (risk-tolerant participants) (*p* > 0.1). Overall, risk-averse participants reduced their work contacts significantly more in 03/2020, and there exists a trend of them maintaining lower levels of work contacts compared to risk-tolerant participants.

**Figure 3:**
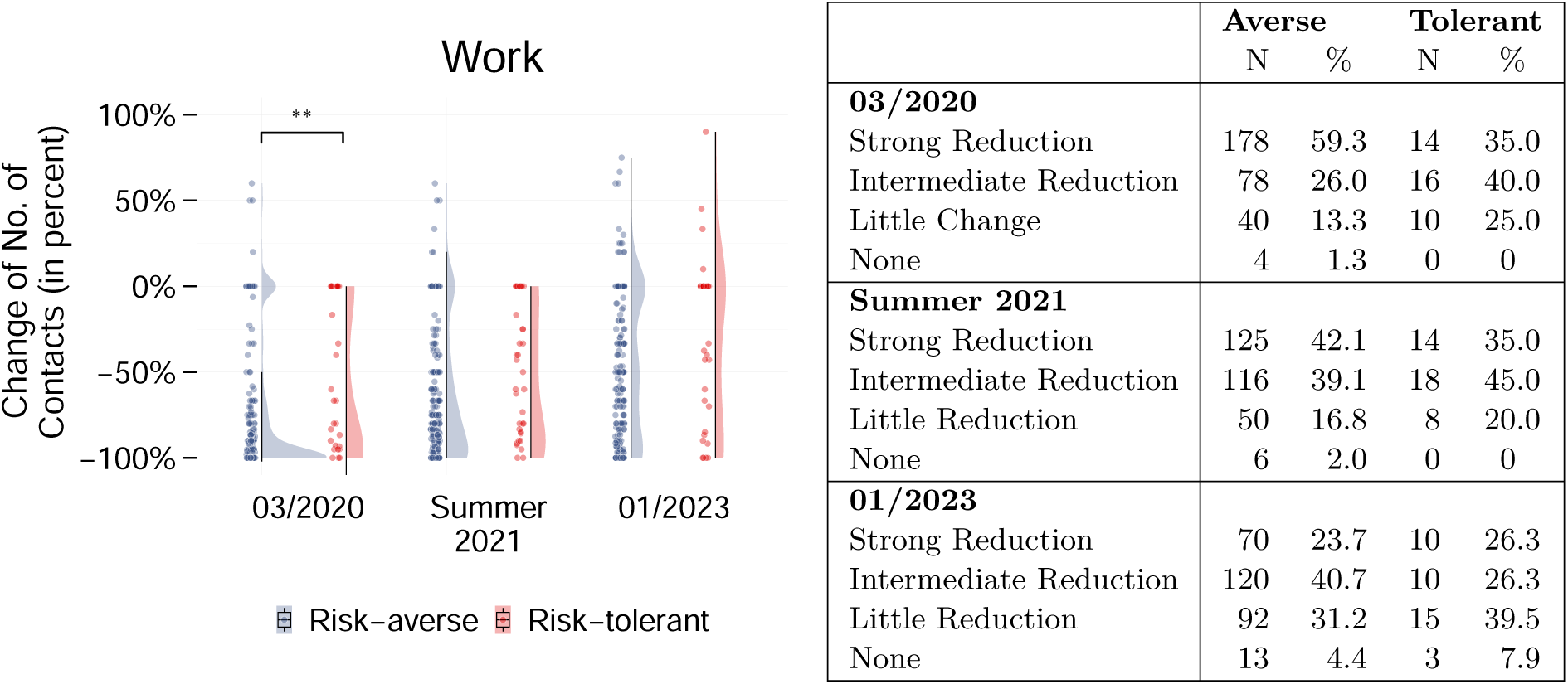
**Left:** Change of number of contacts (in percent, relative to 2019). Stars indicate that the distributions differ at significance levels of \*\**p* < 0.05. On average, both risk perception groups reduced their work contacts for all three time points. In 03/2020, risk-averse participants reduced their work contacts significantly more than risk-tolerant participants. **Right:** Group assignment based on the change of number of contacts (for details, see Subsection Statistical Methods). For all three time points, risk-averse tolerant participants tend to reduce more strongly than risk-tolerant participants.

In the leisure context, risk-averse participants reduced their contacts more strongly and maintained these reductions for longer than risk-tolerant participants. Compared to 2019, risk-averse participants reduced their leisure contacts on average by 77% in 03/2020, while risk-tolerant participants reduced theirs by 53%. Both groups relaxed their leisure contact reductions as time progressed, leading to average reductions of 54% (risk-averse) and 31% (risk-tolerant) in summer 2021, and average reductions of 33% (risk-averse) and 6% (risk-tolerant) in 01/2023 (Fig. 4). The differences between risk perception groups are statistically significant for all three time points (*p* < 0.01). Overall, risk perception consistently influenced leisure contacts, with risk-averse participants reducing more strongly and maintaining the reductions for longer than risk-tolerant participants.

**Figure 4:**
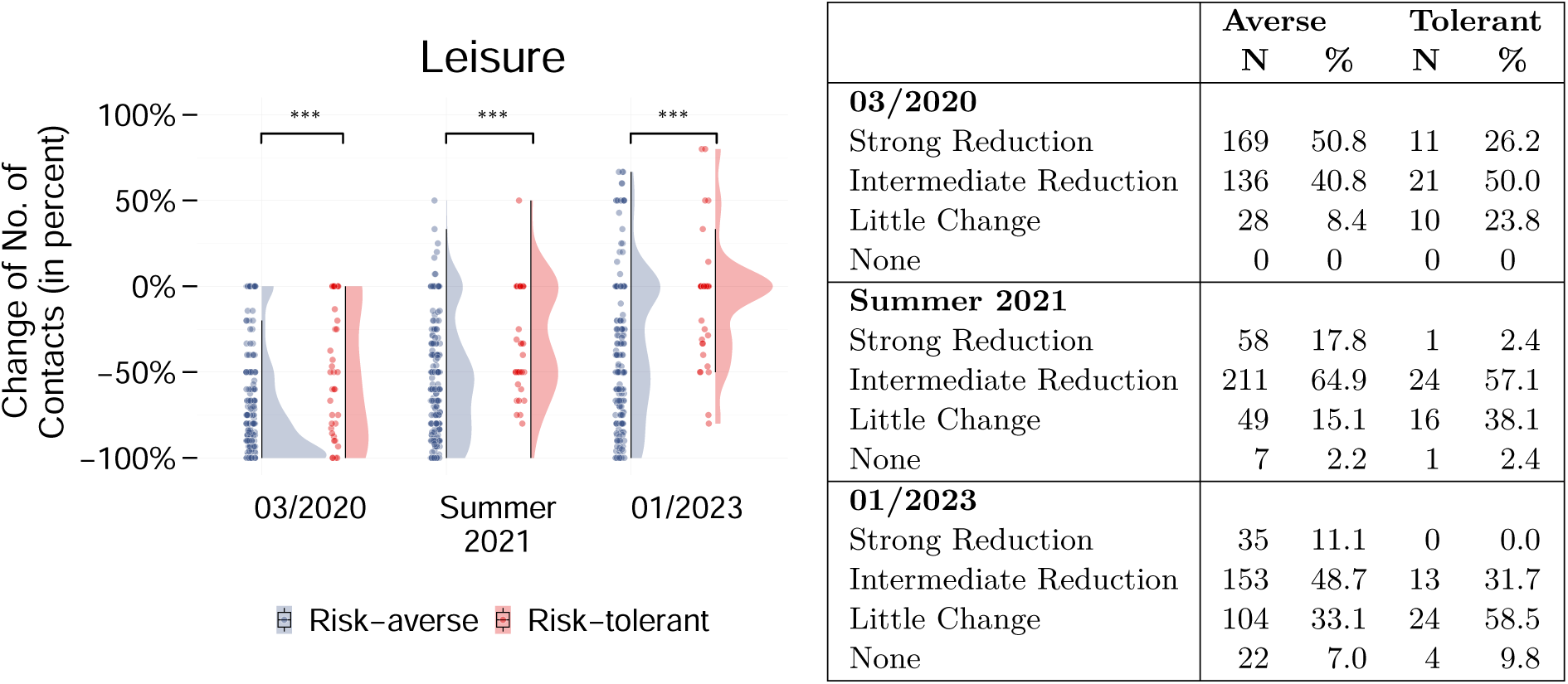
**Left:** Change of number of contacts (in percent, relative to 2019). Stars indicate that the distributions differ at significance levels of \*\*\**p* < 0.01. In 03/2020, more risk-averse than risk-tolerant participants reduced their leisure contacts “strongly”. The share of risk-tolerant participants who “strongly” reduced their leisure contacts was negligible in summer 2021. The majority of risk-tolerant participants had returned to pre-pandemic contact levels in 01/2023. **Right:** Group assignment based on the change of number of contacts (for details, see Subsection Statistical Methods). The difference between risk perception groups already arises in 03/2020 and persists for the two later points in time. Risk-averse participants were more than twice as likely to strongly reduce contacts and half as likely to maintain leisure contacts compared to risk-tolerant participants.

### Correlation of Contact Reductions and Reported COVID-19 Infections

Risk-averse participants reported fewer COVID-19 infections than risk-tolerant participants. Of the risk-averse participants, 39% (95% CI: [33%, 43%]) reported zero infections compared to 32% (95% CI: [19%, 46%]) of risk-tolerant participants. The shares of risk-averse and risk-tolerant participants who reported one COVID-19 infection are comparable (risk-averse: 53%, 95% CI: [47%, 57%], risk-tolerant: 56%, 95% CI: [41,71]). More risk-tolerant than risk-averse participants reported two infections (12% (95% CI: [2%, 21%]) vs 8% (95% CI: [5%, 11%]). The difference in distribution of the number of COVID-19 infections between risk perception groups, however, was not statistically significant (*p* > 0.1) – potentially an artifact of the small number of risk-tolerant participants (Table 2).

**Table 2:** Number of participants per risk perception group.

| Risk Group | N | Percent |
| --- | --- | --- |
| Risk-tolerant | 39 | 11.8 |
| Risk-averse | 292 | 88.2 |
| No Risk Perception Score Available | 372 | - |

Risk-averse participants reported later dates of first infection than risk-tolerant participants, although this difference was not statistically significant. The empirical cumulative distribution functions (ECDF) of the timing of the first infection of risk-averse and risk-tolerant participants are indistinguishable until spring 2021. From spring 2021 onwards, the ECDF for the risk-averse group remained consistently below that of the risk-tolerant group (Fig. 5B). This indicates a trend of risk-averse participants being more successful at avoiding infection than risk-tolerant participants (*p* > 0.1). Overall, the results indicate a correlation between stronger contact reductions and fewer and later COVID-19 infections, but did not reach statistical significance due to the small number of risk-tolerant participants.

**Figure 5:**
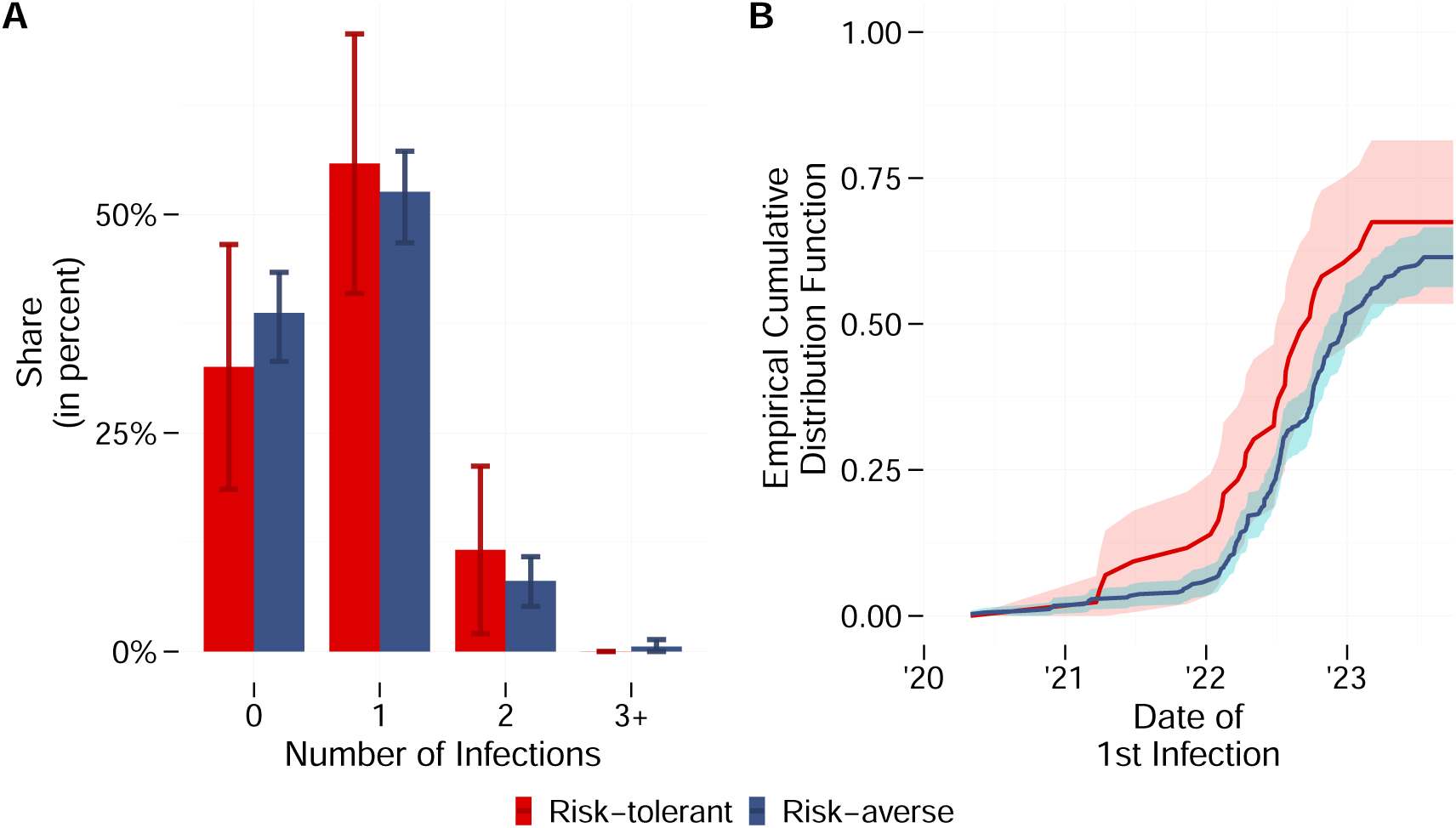
**A.** Share of respondents who reported 0/1/2/3+ infections, differentiated by risk perception score. Error bars represent 95% confidence intervals (see Subsection Statistical Methods for computation and motivation). There exists a trend of risk-tolerant participants reporting more infections than risk-averse participants. **B.** Timing of participants’ (first) infection(s); differentiated by risk perception score. Ribbons represent 95% confidence intervals (see Subsection Statistical Methods for computation and motivation). Risk-averse participants reported fewer infections, and, starting from the spring of 2021, the ECDF of the risk-averse group is constantly below the ECDF of the risk-tolerant group.

### Sub-analyses by Age, Gender, and Comorbidities

18-39 year olds and 40-59 year olds reduced their work contacts more strongly than 60+ year olds (*p* < 0.05, see Supplementary Section Subgroup Analysis by Age Group), but age did not significantly influence leisure contact reductions. Neither gender nor having a COVID-19 relevant comorbidity significantly influenced self-reported contact reductions (*p* > 0.1, Section Subgroup Analysis by Gender, and Section Subgroup Analysis by Comorbidities).

### Contact of Household Members and Closest Contacts

Household members and the closest contacts of participants significantly decreased their work and leisure contacts during the COVID-19 pandemic. Work contact reductions were stronger than leisure contact reductions for both household members and CCs. Similarly to the participants, household members and CCs relaxed their reductions over time. Participants reported the strongest work and leisure contact reductions for themselves, followed by their household members, and finally by their CCs. In the work context, the differences between participants and household members and between participants and CCs are significant for all three time points (*p* < 0.01, see Supplementary Section Contact Reductions of Participant, Household member, Closest contact for details). In the leisure context, the difference between participants and household members and between participants and CCs is statistically significant for all three time points (*p* < 0.01), except for 03/2020: Here, the difference between participants and their household members is not statistically significant (*p* > 0.1). In sum, participants reported stronger reductions for themselves than for their household members and CCs, but the bimodal shape and time-dependency of contact reductions are visible across groups.

### Social Homophily During the COVID-19 Pandemic

Social homophily persisted during the COVID-19 pandemic: The Pearson correlation coefficient of the participants’ and their CCs’ number of contacts is consistently different from zero (p < 0.01). The correlation coefficient is positive in both the work and leisure context and ranges between 0.19 and 0.24 (Table 3). For the risk-averse subgroup, the correlation is larger in magnitude than for the whole sample, ranging between 0.34 and 0.36 in the work context and between 0.29 and 0.35 in the leisure context (Fig. 6). For risk-tolerant participants, on the other hand, the correlation coefficient is not significantly different from zero in the leisure context (*p* > 0.1). As risk-tolerant participants perceived the COVID-19 pandemic as less of a threat, this could indicate that they were not concerned about infection risk due to their second-order contacts. In the work context, risk-tolerant participants’ correlation coefficients are similar to those of risk-averse participants and the overall sample, but are only significantly different from zero at the 10% significance level. In conclusion, the correlation coefficients indicate that the participants preferred to maintain contact with others who reduced their contacts similarly to themselves, providing evidence for the persistence of social homophily during the COVID-19 pandemic.

**Figure 6:**
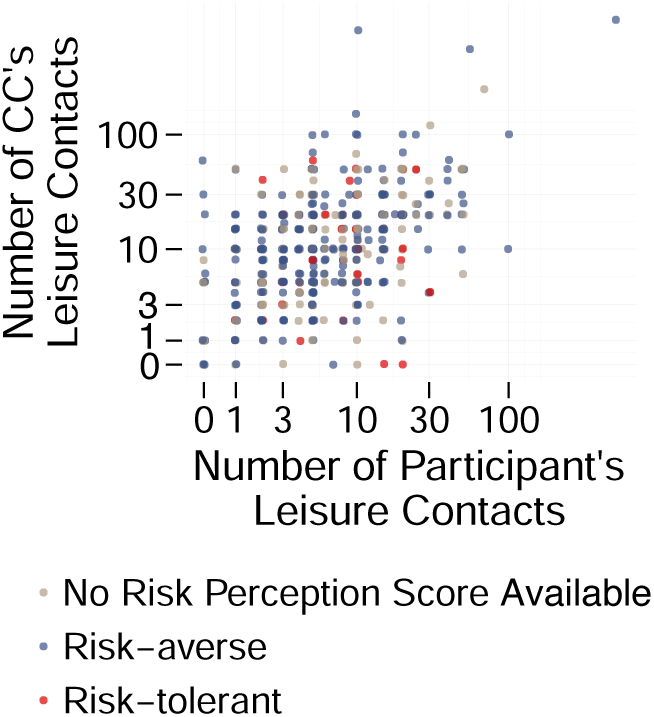
Correlation plot of leisure contacts in 01/2023. Colors depict the different risk perception groups. The number of contacts is depicted on a pseudolog/pseudolog scale to allow the display of 0 contacts.

**Table 3:** Correlation coefficient of participants’ number of contacts and their closest contacts’ number of contacts. Stars indicate that the correlation coefficient is significantly different from zero, with significance levels of \*\*\**p* < 0.01, \*\**p* < 0.05, \**p* < 0.1.

| Time point | All Participants | Risk-Averse Participants | Risk-Tolerant Participants |
| --- | --- | --- | --- |
| **Work** |  |  |  |
| 2019 | 0.21*** | 0.36*** | 0.30* |
| 03/2020 | 0.21*** | 0.35*** | 0.26* |
| Summer 2021 | 0.23*** | 0.34*** | 0.27* |
| 01/2023 | 0.24*** | 0.35*** | 0.28* |
| **Leisure** |  |  |  |
| 2019 | 0.19*** | 0.34*** | −0.07 |
| 03/2020 | 0.21*** | 0.32*** | −0.07 |
| Summer 2021 | 0.22*** | 0.29*** | 0.03 |
| 01/2023 | 0.24*** | 0.35*** | 0.06 |

## Discussion

Our study showed that contact reductions during the COVID-19 pandemic were heterogeneous in three different aspects. First, we quantified that the reductions of work and leisure contacts were both time-dependent and bimodal – with some participants “strongly” reducing, some displaying “little change” in their number of contacts, and the remainder of participants spreading out between the two extremes. Second, we demonstrated that higher risk perception encourages stronger reduction of contacts and, in turn, leads to fewer and later infections. Third, we found evidence that social homophily persisted during the COVID-19 pandemic, especially for risk-averse participants. Considering these heterogeneities may support epidemiological modelers in representing human behavior realistically as well as public health officials and decision makers when implementing measures that aim to reduce contacts.

First, consistent with previous contact studies, we found that both work and leisure contacts were substantially reduced during the pandemic [VHV^+^21, CWG^+^20, WGC^+^23, JCB^+^24]. Notably, we found these reductions to be time-dependent: Contact reductions were relaxed over time, with leisure contacts recover-ing faster than work contacts. Still, by 01/2023, the contacts reported for either context remained below pre-pandemic levels. One potential explanation for the lasting reduction in the work context could be the permanent alteration of office culture: Since the COVID-19 pandemic, more office workers have been able to at least partially work from home and not come into the office every day [MCMW23, Sta21]. This, in turn, reduces their work contacts. In sum, analysis of the mean contact reduction reveals that contacts were most substantially reduced at the beginning of the pandemic, but still remained below pre-pandemic levels by 01/2023.

Examining not only the mean contact reduction but also the full distribution of reductions reveals an important heterogeneity: The distribution of reductions is bimodal, with a peak at either extreme. Some participants reduced their contacts “strongly“, while others showed “little change” in their number of contacts. The shape of this bimodal distribution evolves over time: Typically, participants incrementally relaxed their contact reductions. Still, there exists a noticeable share of participants who moved from the “intermediate” to the “strong” reduction group from 03/2020 to summer 2021. One possible explanation is Germany’s implementation of home office mandates in January 2021. This implementation yielded a reduction of work attendance of the participants, but potentially also of their coworkers. The reduction in attendance then decreased the number of work contacts of the participants. Overall, analysis of the full distribution of contact reductions reveals a bimodal pattern that evolved over time, reflecting heterogeneous behavioral responses. Second, consistent with evidence from other communicable diseases and the health belief model, risk-averse participants who perceived COVID-19 as more dangerous reported stronger contact reductions than risk-tolerant participants [Ros90, Fer07, BM10, LA09, dGCDD22]. This difference in reduction became especially apparent in the leisure context, with risk-averse participants reducing more strongly and upholding these reductions longer. This difference between the two contexts could, on the one hand, indicate a permanent change in office culture. Since the COVID-19 pandemic, remote work has become more common, and online meetings are the norm in many companies [MCMW23, Sta21]. Here, both risk perception groups benefit from the possibility to work from home, and contact reductions are not necessarily a result of the disease spread. On the other hand, the difference may be an indicator of participants’ greater freedom over their leisure contacts. In the leisure context, risk perception can be more easily expressed and acted upon, whereas workplace contacts remain largely dictated by organizational requirements, limiting individuals’ freedom to (not) reduce their work contacts. Differences in contact reductions propagated to differences in infections: Risk-averse participants reported fewer infections, as well as a later date for their first infection. Subanalyses that examined age, gender, and comorbidities did not demonstrate any significant differences in contact reductions. In sum, the findings demonstrate that risk perception was the main driver of contact reductions, with risk-averse participants’ stronger and more sustained reductions translating into fewer infections.

Third, our analysis provides evidence for social homophily during the COVID-19 pandemic. In the work context, the correlation between the participants’ and their closest contacts’ remained constant from 2019 until 01/2023. Differentiated by risk perception score, the correlation coefficient for risk-averse participants was higher than for the risk-tolerant participants. In contrast, in the leisure context, only risk-averse participants displayed a correlation between their own contacts and their closest contact’s number of contacts significantly different from zero. Again, this may be traced back to participants’ freedom over their leisure contacts and lack thereof over their work contacts. These findings demonstrate that social homophily influenced contact patterns during the pandemic, with participants – particularly those who were risk-averse – preferring to maintain relationships with others who reduced their contacts similarly to themselves.

Both our second and our third main results show that risk perception – not demographics – is the main driver of behavior. Recognizing the difference in risk perception may support public health communication and the design of intervention strategies. Tailored messaging that aligns with individual risk perceptions may be more effective than uniform approaches.

While this study has several methodological limitations, our findings are consistent with previous contact studies and contribute valuable insights into pandemic contact behavior. Participants were asked to report their contacts from 2019, 03/2020, summer 2021, and 01/2023 in July and August 2023, potentially introducing recall bias. Especially when reporting contacts from the beginning of the pandemic and when answering the risk perception questions, participants might have been tempted to underestimate their contacts and overestimate their self-protective behaviors to adhere to societal expectations, leading to social desirability bias. Participants were recruited via Twitter: As Twitter users have traditionally been younger, more male, and better educated than the general population, this recruitment method may lead to a biased sample [MLA^+^21]. In our sample, even the most risk-tolerant participants reported reducing their contacts in 03/2020. Our recruitment strategy consequently underrepresented participants who did not consider COVID-19 to be a threat and who, as a result, maintained their pre-pandemic level of contacts, even during peak pandemic periods. Thus, one subpopulation that is of special interest for understanding the state of an outbreak and designing effective public health interventions could not be reached with this recruitment method. Despite these limitations, the distribution of COVID-19 infections and incidence rates observed in our survey are consistent with large-scale studies and official reporting statistics [MNP^+^25]. Our pre-pandemic contact data is comparable to the POLYMOD study for Germany, while our pandemic contact data aligns with previous studies examining contact patterns and activity reductions during this period [TRB^+^21, MPR^+^24, PBN24, PBJ^+^25] (see Supplementary Section Comparison to POLYMOD for details). While these limitations should be considered, they do not diminish the study’s contribution to understanding the intricacies of contact behavior during the COVID-19 pandemic.

Most importantly, our study contributes novel insights by uncovering bimodal patterns of contact reductions and persistent social homophily during the COVID-19 pandemic. These insights reveal substantial heterogeneities in pandemic behavioral response that have been overlooked in previous research. Our results can support epidemiological modelers in representing human behavior more realistically, improving both model accuracy and predictive power [Fer07, FSJ10]. Our findings also have critical implications for public health strategies, as they emphasize the need for non-pharmaceutical interventions that address and exploit these heterogeneities. By tailoring communication strategies and policy measures to these three distinct behavioral groups, public health officials may increase intervention effectiveness while minimizing societal burden. Our findings yield a shift toward targeted public health strategies that account for behavioral heterogeneities, ultimately supporting more effective and sustainable pandemic preparedness for future health crises.

## Data Availability

The survey in the original German is available on OSF (https://doi.org/10.17605/OSF.IO/RTJZU). The anonymized dataset analysed in this study is available on OSF (https://osf.io/7vzgd). Analysis code is available on Zenodo (https://doi.org/10.5281/ZENODO.15783897).

## Availability of Data and Materials

The survey in the original German is available on OSF [FNM^+^23]. The anonymized dataset analysed in this study is available on OSF [CVFN^+^25]. Analysis code is available on Zenodo [Pal25].

## Funding

SP: Ministry of Research and Education (BMBF) Germany (031L0300D, 031L0302A) and TU Berlin

LS: Ministry of Research and Education (BMBF) Germany (031L0300C)

JF: Ministry of Research and Education (BMBF) Germany (031L0300A)

MEK: MEK gratefully acknowledges funding by the The Netherlands Organisation for Health Research and Development (ZonMw) (grant numbers 10710022210003 and 10710062310022).

MM: Ministry of Research and Education (BMBF) Germany (031L0300C)

KN: Ministry of Research and Education (BMBF) Germany (031L0300D, 031L0302A) and TU Berlin

HN: Ministry of Research and Education (BMBF) Germany (031L0300C)

ACV: Ministry of Research and Education (BMBF) Germany (031L0300C)

VP: Ministry of Research and Education (BMBF) Germany (031L0300A), and German Research Foundation (DFG), via the SFB 1528 "Cognition of Interaction".

## Authors contributions

Conceptualization: SP, VP, LS, MEK

Data curation: SP, LS, MM

Formal analysis: SP

Funding acquisition: VP, ACV, KN

Investigation (data collection): LS, MM, JF, MEK, HN, ACV, VP

Methodology: SP, VP, LS, MEK

Project administration: VP

Resources: VP, ACV, KN

Software: SP

Supervision: VP, MEK, ACV, KN

Validation: all

Visualization: SP

Writing - Original Draft: SP, LS, VP

Writing - Review & Editing: all

## Acknowledgements

We thank Jonas Dehning as well as the infoXpand consortium within MONID for the exciting discussions and valuable comments they have provided. Claude AI, Grammarly AI, and LanguageTool were used for grammar checks in the main text, and GitHub Copilot served as a coding assistant. The authors assume full responsibility for the final content of the article.

## Supplementary material

### Participant Characteristics

The online survey was started by 867 and completed by 398 participants. The majority of survey participants was between 40 and 59 years old, less than one percent were over the age of 80 (Supplementary Figure 1). Around 54% of the participants reported their gender as female, 45% as male, and *<* 1% as diverse. The mean household size in the survey was 2.7. A detailed analysis of the demographic composition of the sample can be found in our previous publication [MNP^+^25].

**Supplementary Figure 1:**
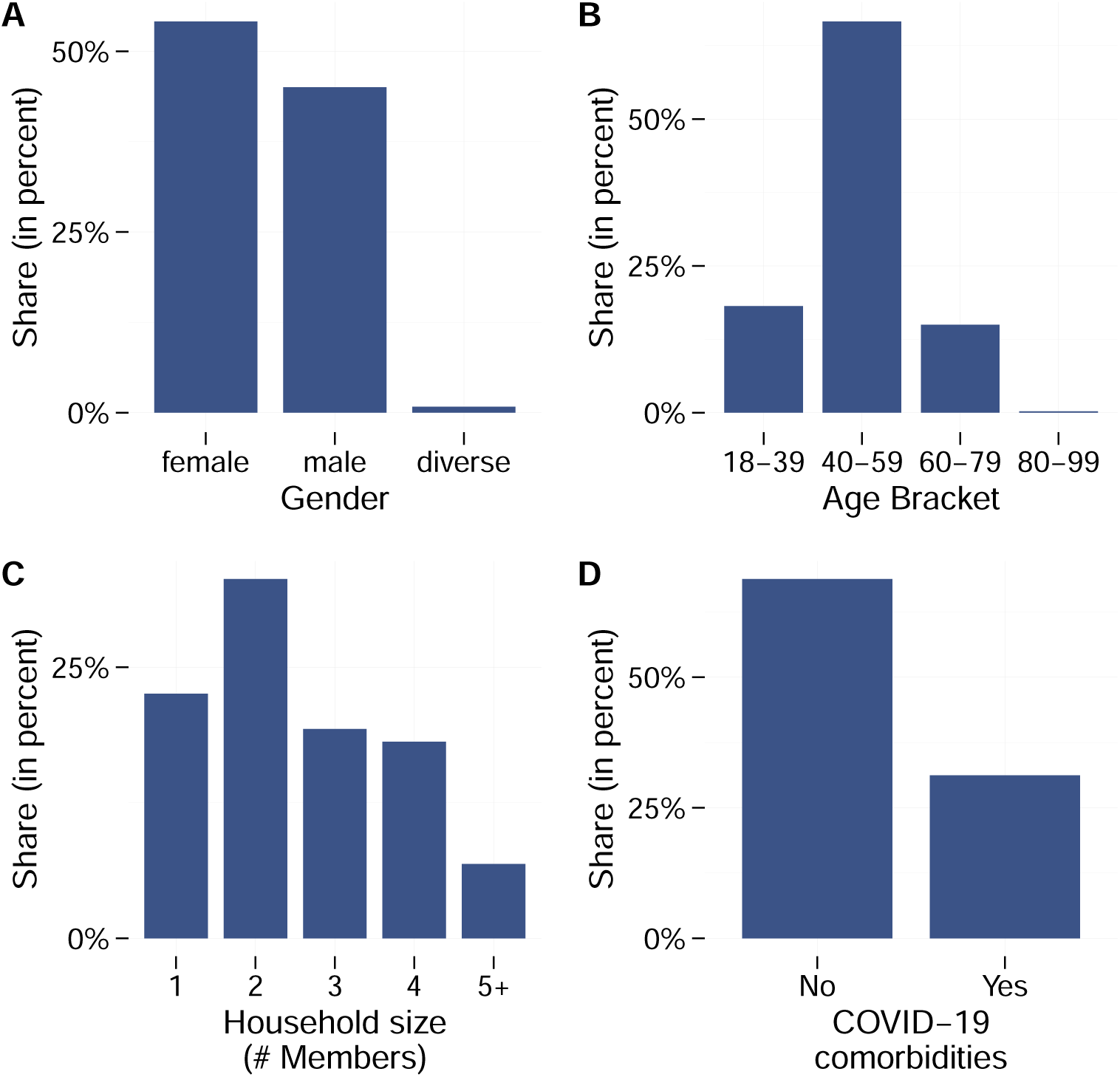
Participants’ demographic characteristics. **A.** Gender distribution. 54% of participants reported their gender as female. **B.** Age distribution. Around 66% of participants were between 40 and 50 years old. **C.** Household size distribution. 1-Person-Households are undersampled. **D.** Comorbidities distribution. Around 31% reported a COVID-19 relevant comorbidity.

### Pre-pandemic Contact Data

For 2019, participants reported more work than leisure contacts. On average, participants reported 37.4 weekly work contacts (median: 20, IQR: 10-40) and 14.4 leisure contacts (median: 10, IQR: 5-20). The distribution of participants’ work contacts is heavy-tailed and unimodal, while the distribution of leisure contacts is also heavy-tailed, but displays three bumps around 5, 10, and 20 (Supplementary Figure 2). Overall, work contacts dominated participants’ pre-pandemic contact behavior, with around 2.5 times more contacts reported in the work context than in leisure settings.

**Supplementary Figure 2:**
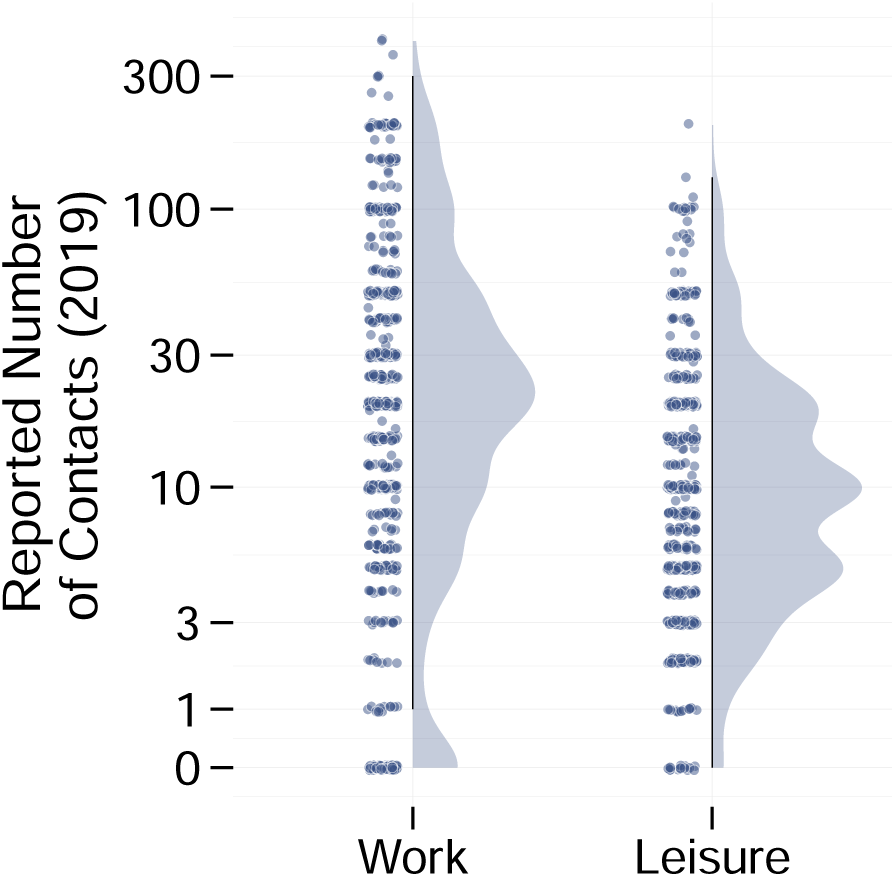
Distribution of survey participants’ number of weekly work (left) and leisure (right) contacts in 2019. Participants reported more work than leisure contacts.

### Comparison to POLYMOD

In contrast to our survey, the POLYMOD study recruited underage participants, but the remaining participant characteristics are comparable. The POLYMOD study surveyed 1,341 German participants, about 1.5 times more than our survey (Supplementary Table 1). Just like our survey, the POLYMOD study failed to recruit the elderly. They did, however, survey underage participants. The gender distribution and household sizes in the POLYMOD sample are comparable to our survey sample.

The comparability of pre-pandemic contact numbers according to our survey and the POLYMOD study is limited due to the different study designs. First, the definition of “contact” differed slightly in the two studies: In the POLYMOD study, a contact was characterized as either a physical contact involving skin-to-skin contact (e.g. handshake or kiss), or a nonphysical contact consisting of a two-way conversation of at least three words in the physical presence of another person but without skin-to-skin contact. In our survey, we defined a “contact” as any situation in which one person comes closer than two meters to another person for at least 15 minutes. Second, we asked for the weekly number of contacts, whereas the POLYMOD study surveyed the number of contacts on a single day. Third, our online survey was conducted in July and August 2023 and retrospectively collected the number of contacts, while POLYMOD asked participants to report their contacts for the previous day. Consequently, in POLYMOD, the mean number of work contacts is 2.5 (median: 0, IQR: 0-4), and the mean number of leisure contacts is 2.3 (median: 2, IQR: 0-3) (Supplementary Figure 3). Again, comparability is limited and the difference in definition of contact cannot be overcome, but to naively transform the weekly number of the survey to daily number of contacts, we divided them by seven and rounded down. Consequently, we obtain a mean number of daily work contacts of 5 (median: 2, IQR: 1-5) and a mean number of daily leisure contacts of 2 (median: 1, IQR: 0-2). Following this transformation, the daily number of work contacts in our study and in the POLYMOD study are of the same magnitude. Still, in comparison to the POLYMOD study, the number of survey participants who reported zero daily work contacts is significantly smaller. Daily leisure contacts are comparable. Overall, when transformed to daily contacts, our data shows comparable contact patterns to those found in POLYMOD despite the different study designs.

**Supplementary Table 1:** Participant characteristics in our survey compared to the POLYMOD study. Our survey did not include underage participants and oversampled 40-59-year-olds. The gender and household distributions are comparable in both studies. *The POLYMOD study only allowed participants to report their gender as either female or male.

|  | **SURVEY** |  | **POLYMOD** |  |
| --- | --- | --- | --- | --- |
|  | **N** | **%** | **N** | **%** |
| **Age category** |  |  |  |  |
| 0-17 | 0 | 0.0 | 361 | 27.9 |
| 18-39 | 155 | 18.1 | 369 | 28.5 |
| 40-59 | 569 | 66.7 | 347 | 26.8 |
| 60-79 | 128 | 15.0 | 205 | 15.8 |
| 80+ | 2 | 0.2 | 13 | 1.0 |
| Missing | 13 | — | 46 | — |
| **Sex of participants** |  |  |  |  |
| Female | 464 | 54.2 | 722 | 55.4 |
| Male | 386 | 45.0 | 581 | 44.6 |
| Diverse | 7 | 0.8 | NA* | NA |
| Missing | 19 | — | 38 | — |
| **Household size** |  |  |  |  |
| 1 | 194 | 22.6 | 250 | 18.6 |
| 2 | 285 | 33.1 | 411 | 30.6 |
| 3 | 166 | 19.3 | 339 | 25.3 |
| 4+ | 215 | 25.0 | 341 | 25.4 |
| Missing | 7 | — | 0 | — |

**Supplementary Figure 3:**
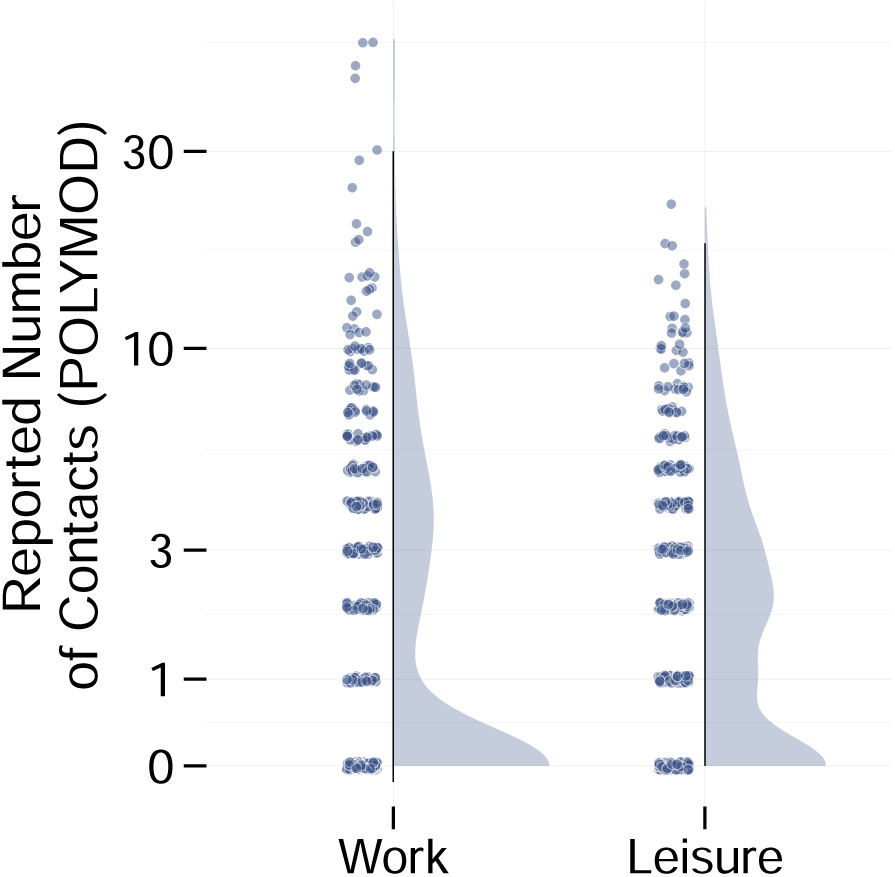
Distribution of Germany POLYMOD participants’ number of daily work and leisure contacts. Work raincloud plot only includes participants who reported that they were working at the time of the study. The median (IQR) number of work contacts is 0 (0-4) and the median (IQR) number of leisure contacts is 2 (0-3).

### Auxiliary Information on Time Points of Interest

The survey was conducted in July and August 2023. Consequently, absolute contact numbers for 2019, 03/2020, summer 2021, and 01/2023 were collected retrospectively. To help participants recall their contact behavior at these time points, auxiliary information was provided on the then-current state of the COVID-19 pandemic (Supplementary Table 2).

**Supplementary Table 2:** Additional contextual information provided to the online survey participants when they were surveyed about their contacts at the four specific time points.

| Time point description (original German) | Time point description (English translation) |
| --- | --- |
| *Im Jahr 2019*, vor der Pandemie: Hier geht es um einen Zeitpunkt vor der Pandemie, z.B. eine typische Arbeitswoche im Jahr 2019. | *2019*: A point in time before the pandemic, for example, a typical working week in 2019. |
| *Ende März 2020*: Die erste Coronawelle breitet sich aus, die Schulen werden geschlossen, der erste Lockdown wirdverordnet, die Pandemie beginnt. Die Bilder aus Bergamo und New York gehen um die Welt. | *End of March 2020*: The first coronavirus wave spreads, schools are closed, the first lockdown is imposed and the pandemic begins. The images from Bergamo and New York go around the world. |
| *Sommer 2021*: Die Impfkampagne läuft an. Die Delta-Variante tritt auf. Die Maßnahmen sind abhängig von der lokalen Inzidenz (Grenzwerte 35 und 50) sowie vom persönlichen Impfstatus. Die Inzidenzen bleiben eher niedrig. Die Fussball-EM findet in vielen Großstädten-verteilt in ganz Europa statt. | *Summer 2021*: The vaccination campaign starts. The Delta variant occurs. The measures depend on the local incidence (threshold values of 35 and 50) and the personal vaccination status. Incidence rates remain rather low. The European Football Championship takes place in many major cities across Europe. |
| *Januar 2023*: Viele Maßnahmen wurden eingestellt. Es gibt einen bivalenten Impfstoff. In ÖPNV und der Bahn gilt vielerorts noch die Maskenpflicht. | *January 2023*: Many measures have been discontinued. A bivalent vaccine is available. Masks are still compulsory on public transport and trains at many locations. |

### Survey Items on Attitudes Related to Covid-19

Nine of the survey items surveyed participants regarding their attitudes related to COVID-19 at the end of March 2020. Participants were asked to report their attitude compared to an “average person” (Supplementary Table 3 for survey items). Possible answers ranged from “a lot less” to “a lot more”, but also included the options “does not apply” and “not specified” (Supplementary Table 4). For the analysis, the options “does not apply” and “not specified” were excluded, yielding a 7-point Likert scale.

**Supplementary Table 3:**
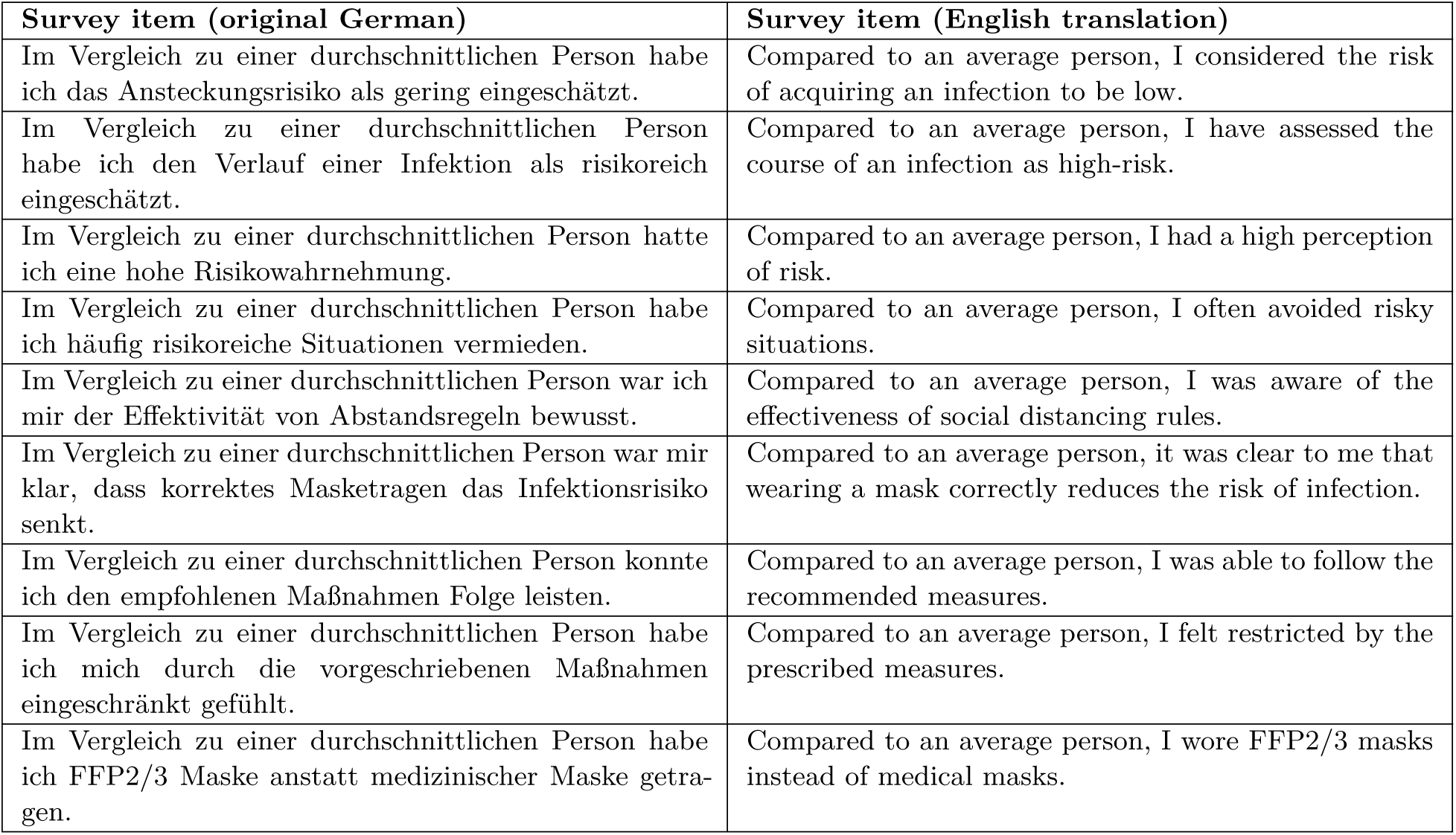
Survey items on attitudes related to COVID-19. The beginning of the corresponding survey section states that these items refer to the end of March 2020 and asked participants to report their own attitudes in comparison to an average person.

**Supplementary Table 4:**
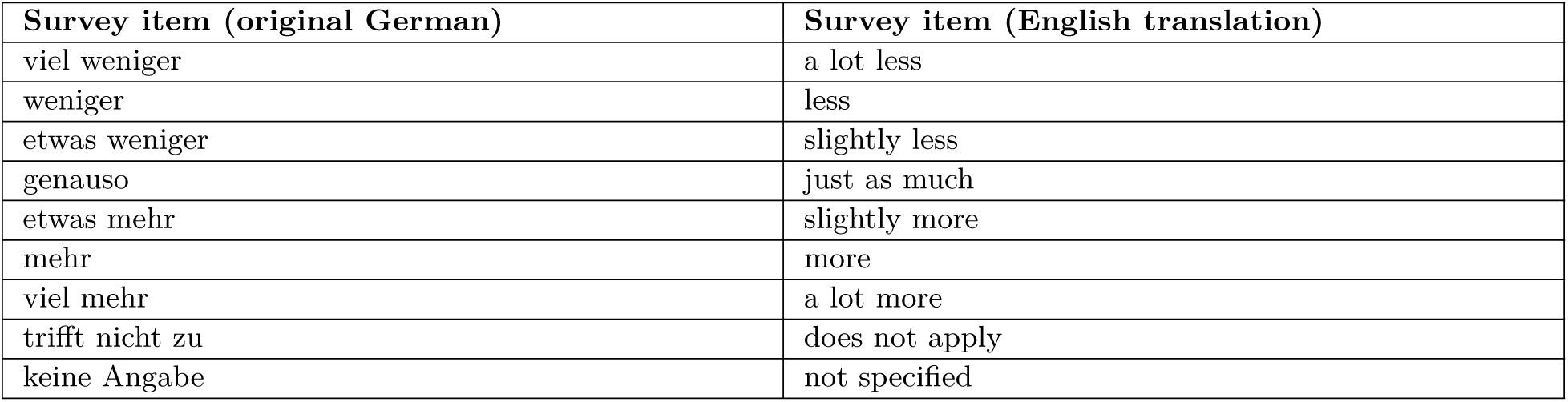
Answer options for items on attitudes related to COVID-19. “Does not apply” and “not specified” were excluded from the analysis, yielding a 7-point Likert scale.

| Survey item (original German) | Survey item (English translation) |
| --- | --- |
| viel weniger | a lot less |
| weniger | less |
| etwas weniger | slightly less |
| genauso | just as much |
| etwas mehr | slightly more |
| mehr | more |
| viel mehr | a lot more |
| trifft nicht zu | does not apply |
| keine Angabe | not specified |

### Bayesian Fits

We used Bayesian modeling to quantify the shape of the distribution of change of number of work or leisure contacts at each time point. Gaussian mixture models were used, fitting one to four (half-)normal distributions to the data using Markov chain Monte Carlo. Means of the distribution were assumed to be fixed, variances and weights of the distributions were estimated. The following four models were fitted to the data:

- **One distribution**: Half-normal distribution shifted to *−*100%, variance estimated.
- **Two distributions**: Half-normal distribution shifted to *−*100%, normal distribution with a mean of 0%. Variance and weight of distributions estimated.
- **Three distributions**: Half-normal distribution shifted to *−*100%, normal distribution with a mean of 0%, normal distribution with a mean of *−*50%. Variance and weight of distributions estimated.
- **Four distributions**: Half-normal distribution shifted to *−*100%, normal distribution with a mean of 0%, normal distribution with a mean of *−*50%, normal distribution with a mean of 50%. Variance and weight of distributions estimated.

For each context and each point in time, we used leave-one-out cross validation to compare the different fits. With the exception of the leisure context in 01/2023, the cross validation always favored three distributions (see Supplementary Table 5). For each context and each time point, we used the posterior mean estimates to estimate group – “strong reduction”, “intermediate reduction”, and “little change” – membership probabilities. Based on these probabilities, participants were assigned to the three groups (see Supplementary Figure 4 for exemplary model results for work 03/2020).

**Supplementary Figure 4:**
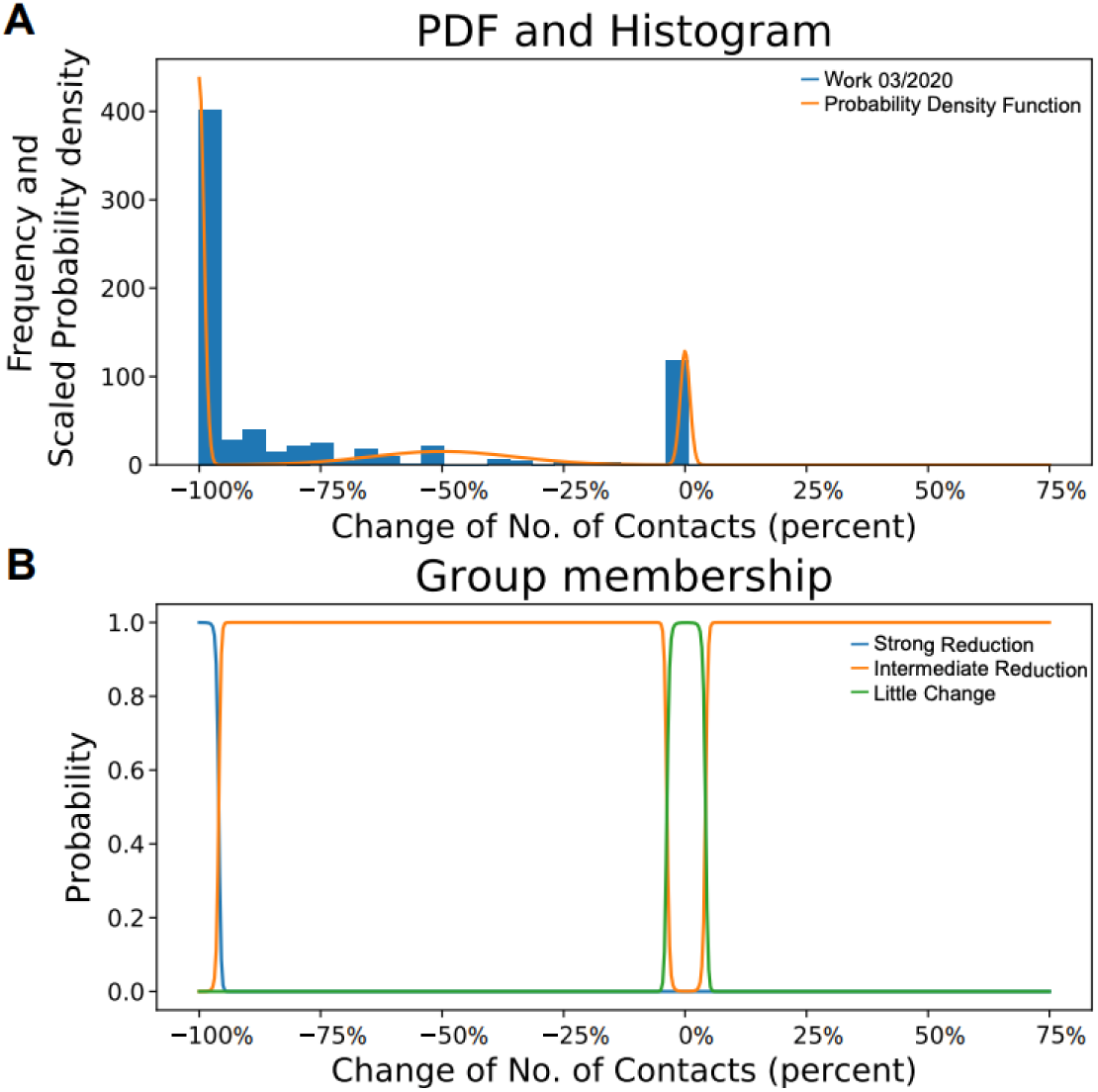
Exemplary depiction of Bayesian model results for the work context in 03/2020. **A.** Histogram depicting the distribution of the change of number of contacts (blue). Bayesian models were fit to this data and the scaled probability density function of the three distributions model is depicted in yellow **B.**. Probability of assignment to groups “strong reduction” (blue), “intermediate reduction” (yellow), and “little change” (green).

**Supplementary Table 5:**
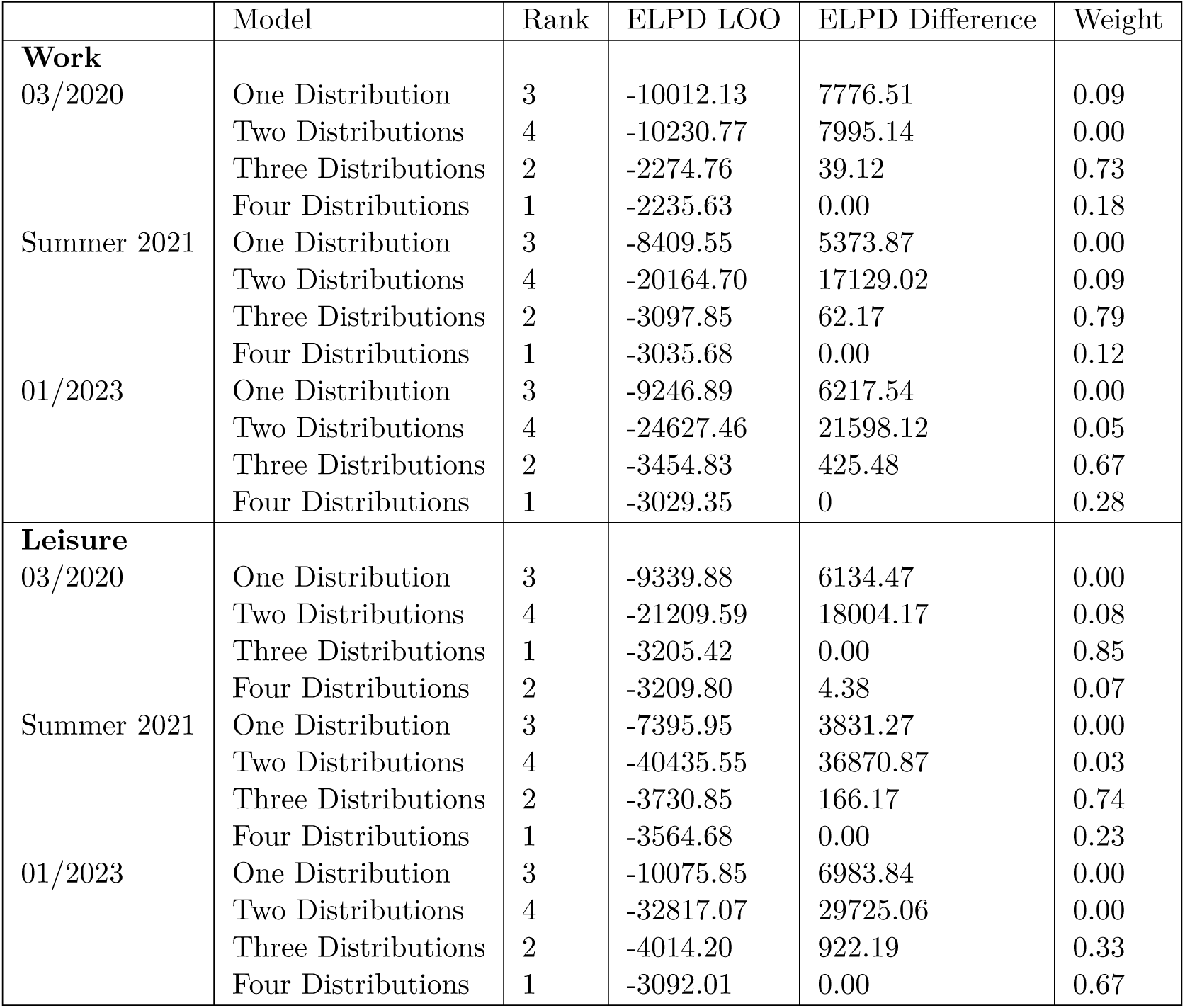
Model comparison results. Leave-one-out cross-validation, an estimate of the out-of-sample predictive fit, was used for model comparison. The weights provided in column six may loosely be interpreted as the probability of each model being true. With the exception of the leisure context in 01/2023, the largest weight is always placed on the model fitting three distributions.

### Pre-Pandemic Contact Data Differentiated by Risk-Perception Score

The two risk perception groups reported comparable work and leisure contacts for 2019. For risk-tolerant participants, the mean of weekly work contacts was 33 (median: 20, IQR: 10-50), while for risk-averse participants the mean was 37 (median: 20, IQR: 8-40). Similarly, the mean number of leisure contacts was 11 for the risk-tolerant participants (mean: 10, IQR: 5-15) and 14 for the risk-averse participants (IQR: 5-20) (Supplementary Figure 5A). The difference in distribution was neither statistically significant for the work nor for the leisure context (*p* > 0.1, see Supplementary Section Kolmogorov-Smirnov tests). The two risk perception groups display no apparent difference in their pre-pandemic sociability.

**Supplementary Figure 5:**
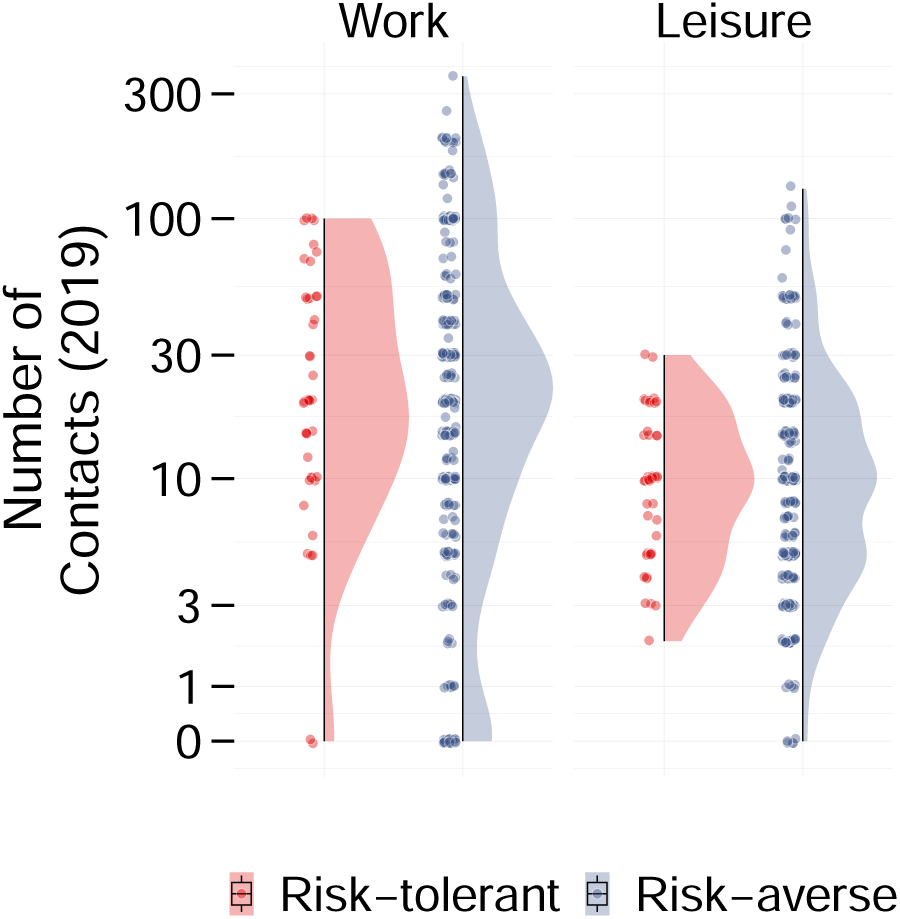
**A.** Distribution of participants number of work (left) and leisure (right) contacts in 2019 differentiated by risk perception group. For both groups and both contexts, the distribution of the number of contacts is unimodal. There exists no significant difference in the distribution of the number of contacts between the two groups.

### Accuracy of Participants’ Reported Contact Numbers

Our validation approach to examine participants’ accuracy in reporting their CCs’ behavior yielded limited data, with only nine analyzable pairs allowing for qualitative assessment. To examine how accurately participants reported their closest contacts’ number of contacts, we invited them to forward the survey to the CCs they had been reporting on throughout the survey. Only 22 participants followed this invitation and forwarded the survey to their CC. This yielded 22 pairs of “original” and “referred” participants. Of these 22 pairs, we only analyzed the pairs for which a) the original participant forwarded the survey to exactly one contact and b) both the “original” and the “referred” participant reported that they had not changed their CC during the COVID-19 pandemic. This promotes that the closest contact described in the survey is the same person who received the forwarded survey. This filtering process resulted in nine pairs and only allows qualitative comparison. Overall, only a fraction of participants forwarded the survey to their CC, allowing only a qualitative assesment of the participants’ accuracy.

Participants reported the CCs’ household members and leisure contacts reasonably well, but differed in their ability to report the CCs’ work contacts. The “original” participants, with the exception of a deviation of five for the 5th pair and a deviation of one for the 9th pair, reported their CC’s number of household members accurately (Supplementary Figure 6). Apart from the 3rd pair, the “original” participants reported the CCs’ number of leisure contacts accurately (Supplementary Figure 8). The “original” participants of the 2nd, 5th, 7th, and eight 8th pair, estimated the number of the CCs’ work contacts reasonably well, while the “original” participants of the remaining pairs (1st, 3rd, 4th, 6th, and 9th) pair misjudged the number of work contacts of the CCs (Supplementary Figure 7). Interestingly, with the exception of the 9th pair, all “original” participants overestimated the number of work contacts of their CC. Overall, we note that participants accurately reported the CCs’ leisure contacts, but not all participants succeeded at reporting the CCs’ work contacts.

**Supplementary Figure 6:**
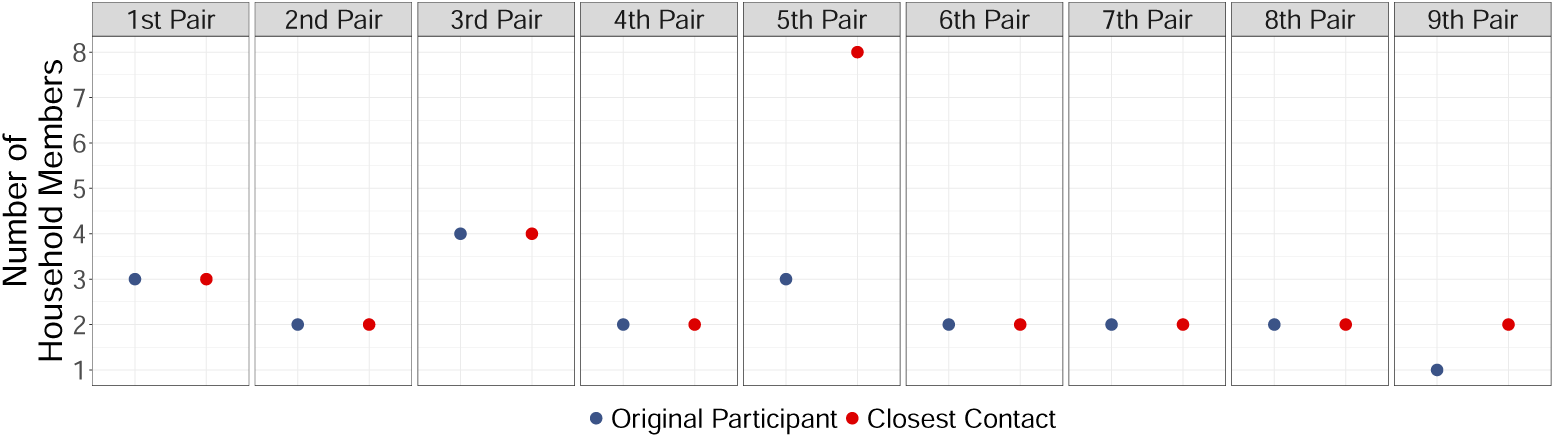
Number of household members the original participants reported for their CC (blue) vs number of household members the CC themselves reported (red).

**Supplementary Figure 7:**
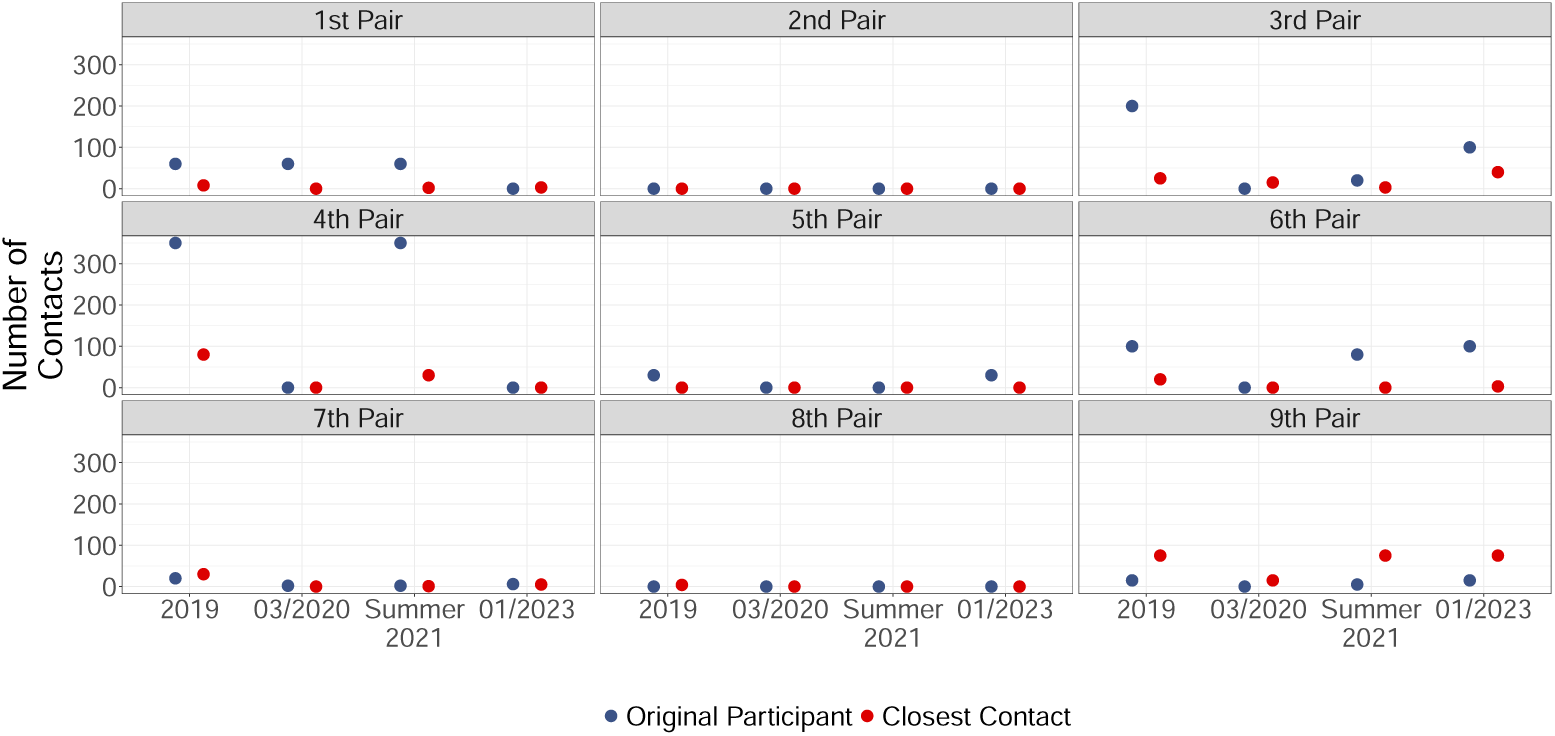
Number of work contacts the original participants reported for their CC (blue) vs number of work contacts the CC themselves reported (red).

**Supplementary Figure 8:**
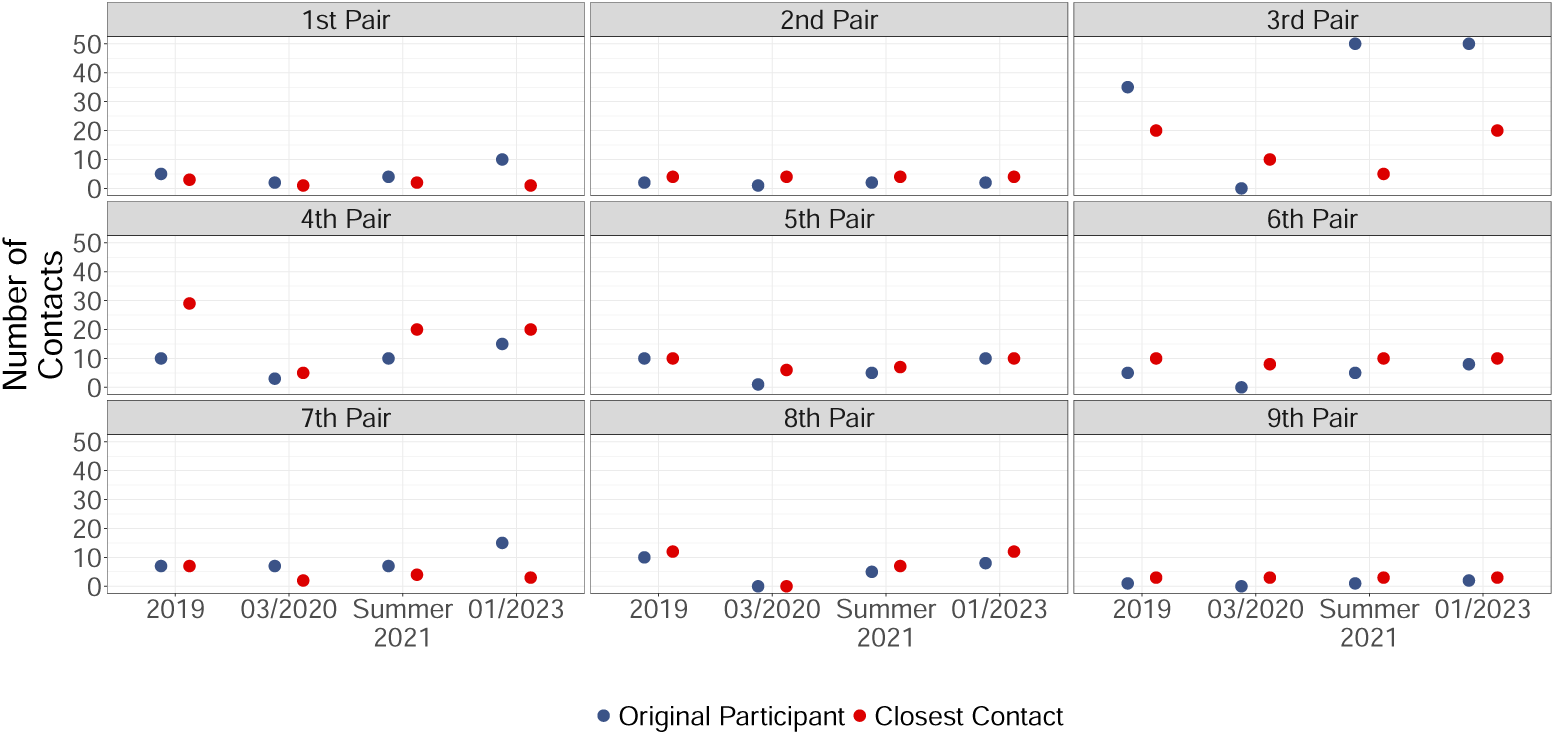
Number of leisure contacts the original participants reported for their CC (blue) vs number of leisure contacts the CC themselves reported (red).

### Contact Reductions of Participant, Household member, Closest contact

The survey asked participants to report

(a) their own contacts,
(b) their household members’ contacts (only if the participant did not live in a one-person-household),
(c) their closest non-household contact’s contacts,
(d) if they reported having changed their closest contact during the COVID-19 pandemic, their novel closest contact’s contacts.

**Supplementary Figure 9:**
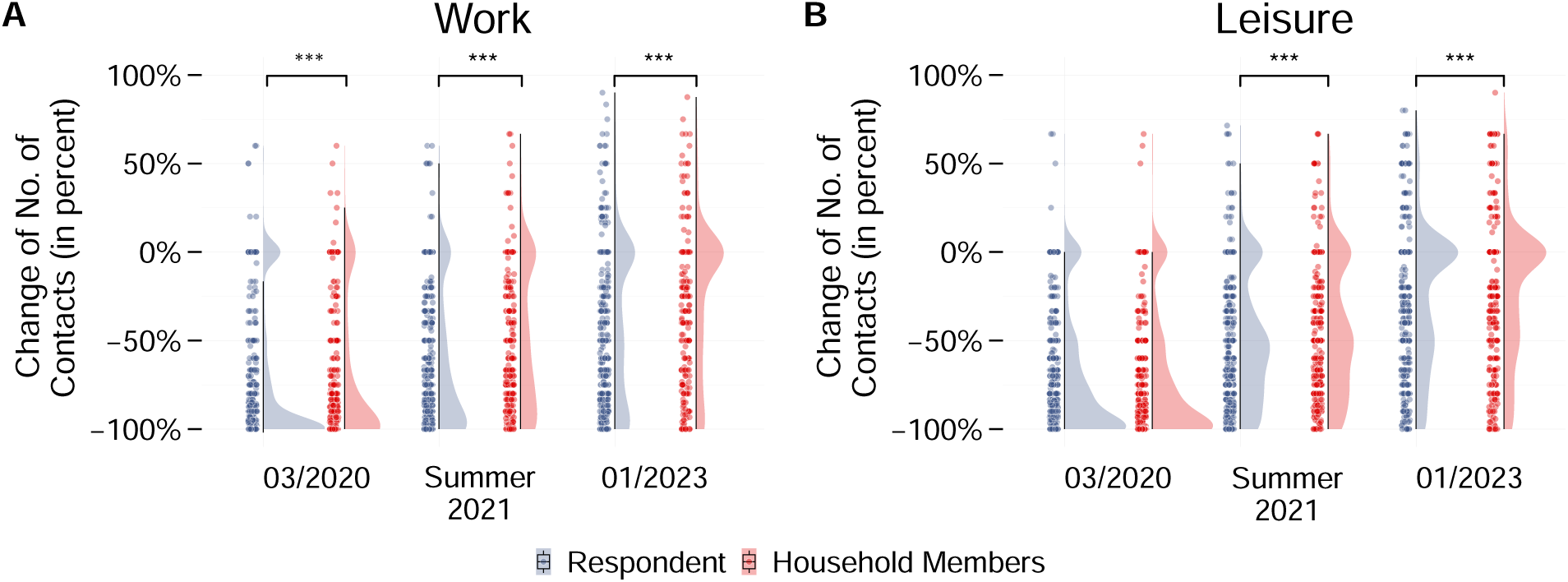
Change of number of contacts (in percent, relative to 2019) of participants vs of household members. **A. Relative number of work contacts.** Analogously to the participants, household members reduced their work contacts at all three time points. For all three time points, fewer household members than participants “strongly” reduced their work contacts. **B. Relative number of leisure contacts.** Overall, the shape of the distributions are similar for participants and household members.

For 03/2020, summer 2021, and 01/2023, participants reported the largest average work contact reduction for themselves, followed by their household members, and finally by their closest contact (CC): In March 03/2020, participants reported an average reduction of 75% for themselves, of 71% for their household members, and of 69% for their CC. These reductions were relaxed from 03/2020 to summer 2021, leading to an average reduction of 64% for the participants, of 49% for the household members, and of 46% for the CCs. Reductions were futher relaxed such that for 01/2023, participants reported an average work contact reduction of 42% for themselves, of 27% for their household members, and of 21% for their CC (Supplementary Figure 9 A and Supplementary Figure 10 A). These differences in mean reductions are mirrored by the differences of the Bayesian reduction model: For all three time points, the share of “strong reduction” is largest for the participants, second largest for the household members, and smallest for the CCs. Complementary, the share of “little change” is always largest for the CCs, followed by household members, and finally by the participants (Supplementary Table 6). Differences in the distribution of change of work contacts between (a) participants and their household members and (b) participants and their CCs are statistically significant for all three time points (*p* < 0.001, Supplementary Table 8). Overall, participants reported the strongest work contact reductions for themselves, but the overall work behavior patterns – with a bimodal distribution and incrementally relaxing reductions – are visible for the household members and CCs, too.

**Supplementary Table 6:**
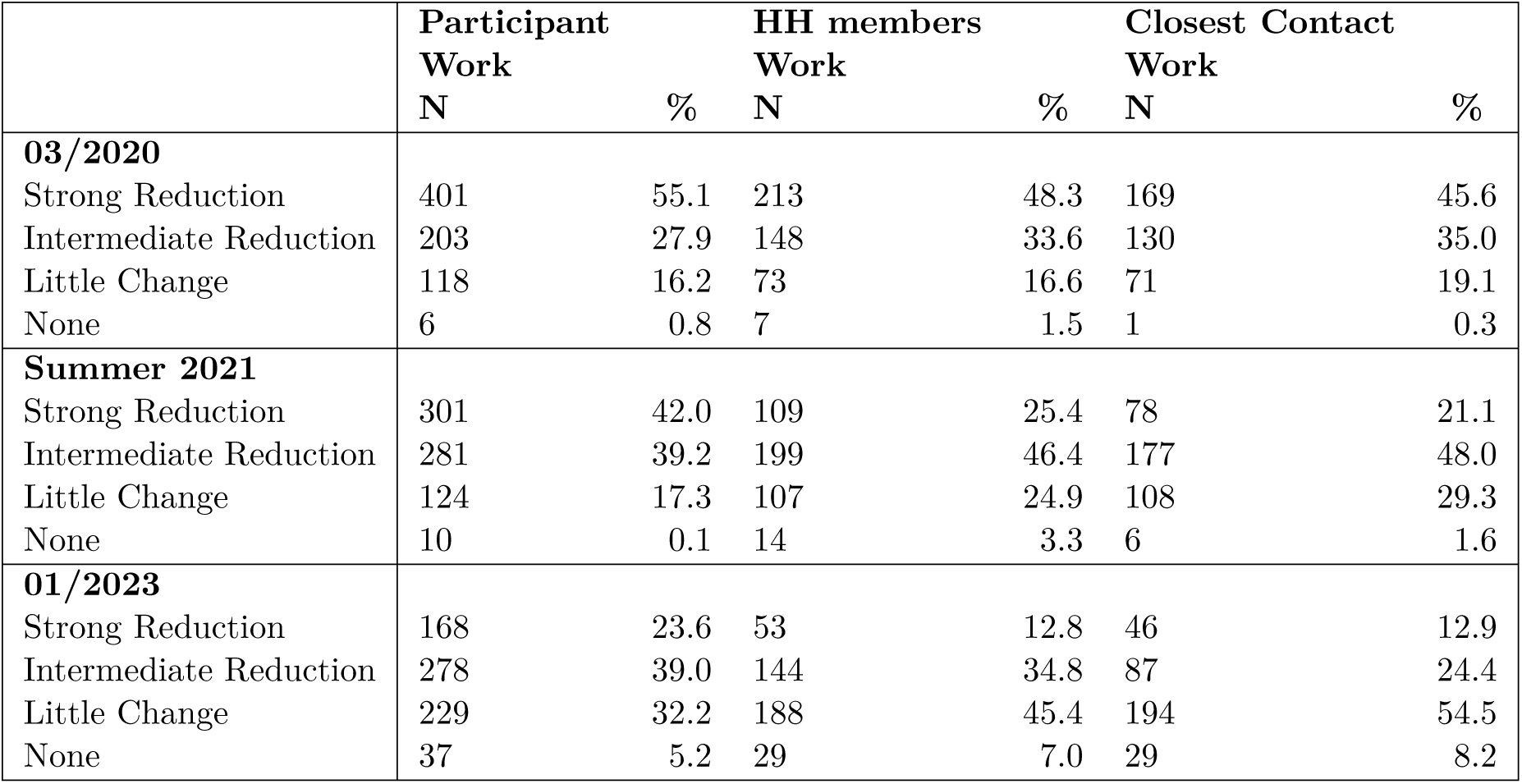
Group assignment based on the change of no. of contacts (for details see Subsection Statistical Methods). Across all three points in time, participants reported themselves to reduce their work contacts more than their household members who, in turn, reduced their work contacts more than the participants’ CCs.

|  | **Participant Work** |  | **HH members Work** |  | **Closest Contact Work** |  |
| --- | --- | --- | --- | --- | --- | --- |
|  | **N** | **%** | **N** | **%** | **N** | **%** |
| **03/2020** |  |  |  |  |  |  |
| Strong Reduction | 401 | 55.1 | 213 | 48.3 | 169 | 45.6 |
| Intermediate Reduction | 203 | 27.9 | 148 | 33.6 | 130 | 35.0 |
| Little Change | 118 | 16.2 | 73 | 16.6 | 71 | 19.1 |
| None | 6 | 0.8 | 7 | 1.5 | 1 | 0.3 |
| **Summer 2021** |  |  |  |  |  |  |
| Strong Reduction | 301 | 42.0 | 109 | 25.4 | 78 | 21.1 |
| Intermediate Reduction | 281 | 39.2 | 199 | 46.4 | 177 | 48.0 |
| Little Change | 124 | 17.3 | 107 | 24.9 | 108 | 29.3 |
| None | 10 | 0.1 | 14 | 3.3 | 6 | 1.6 |
| **01/2023** |  |  |  |  |  |  |
| Strong Reduction | 168 | 23.6 | 53 | 12.8 | 46 | 12.9 |
| Intermediate Reduction | 278 | 39.0 | 144 | 34.8 | 87 | 24.4 |
| Little Change | 229 | 32.2 | 188 | 45.4 | 194 | 54.5 |
| None | 37 | 5.2 | 29 | 7.0 | 29 | 8.2 |

In the leisure context and with the exception of 03/2020, participants again report the largest average reductions for themselves, followed by their household members and their CCs: In 03/2020, the mean leisure contact reduction was 72% for the participants, 73% for the household members, and 69% for the CCs. Participants, household members and CCs relaxed their reduction of leisure contacts from 03/2020 to summer 2021, leading to mean reductions of 51% (participants), 44% (household members), and 36% (CCs). Finally, for 01/2023, participants reported a mean leisure reduction of 28% for themselves, of 23% for their household members and of 9% for their CCs (Supplementary Figure 9 B and Supplementary Figure 10 B). The Bayesian reduction model mostly determines the largest share of “strong reduction” for the participants themselves, closely followed by their household members, and then finally by the CCs (Supplementary Table 7). The only exception is 03/2020, when the share of household members who “strongly” reduced leisure contacts is larger than the share of participants who “strongly” reduced leisure contacts. The distributions of leisure contact reductions of participants and household members are not statistically different in 03/2020 (*p* > 0.1, Supplementary Table 7). For the two subsequent time points, however, and for all three time points for the participants and the CCs, the difference is statistically significant (*p* < 0.01). In sum, the behavior of the household members and CCs mirrors the behavior of the participants, with strong leisure reductions during the early phase of the COVID-19 pandemic and incremental relaxations.

**Supplementary Figure 10:**
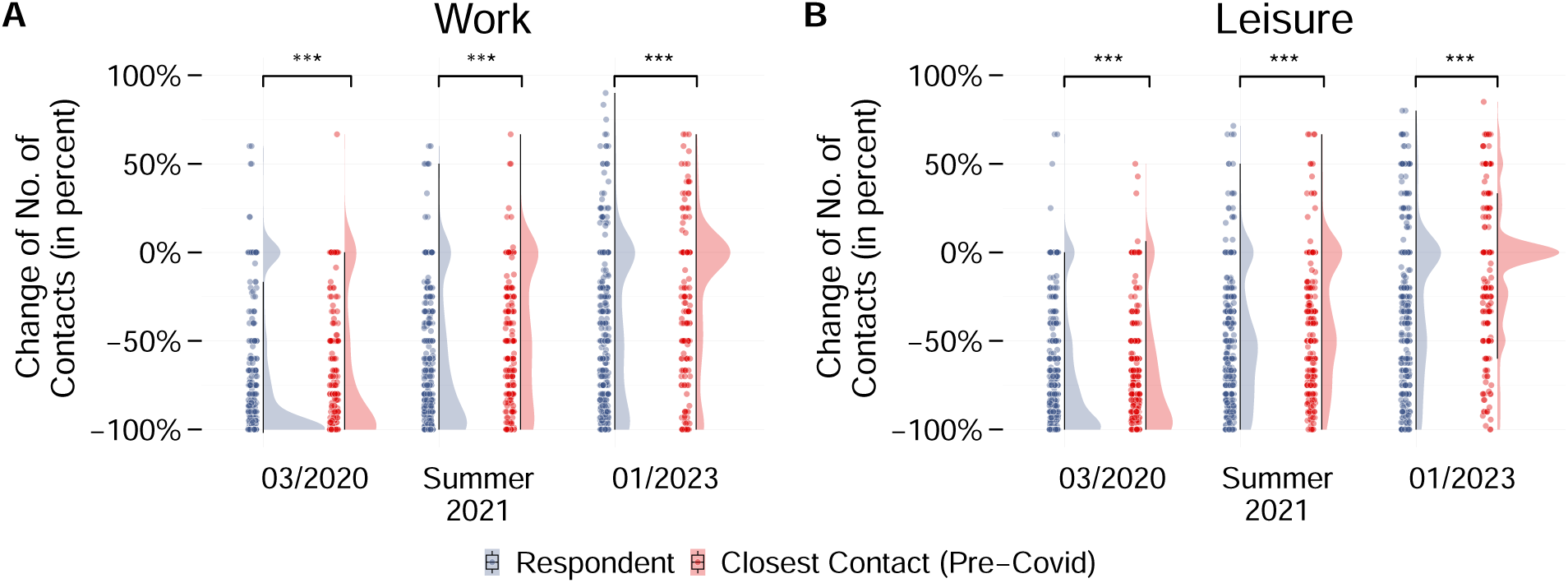
Change of number of contacts (in percent, relative to 2019) of participants vs of their closest contacts. As a percentage increase from zero cannot be computed, individuals who reported zero work/leisure contacts in 2019, were excluded from the corresponding context. **A.** In comparison to the share of participants who completely cut their work c,

**Supplementary Table 7:**
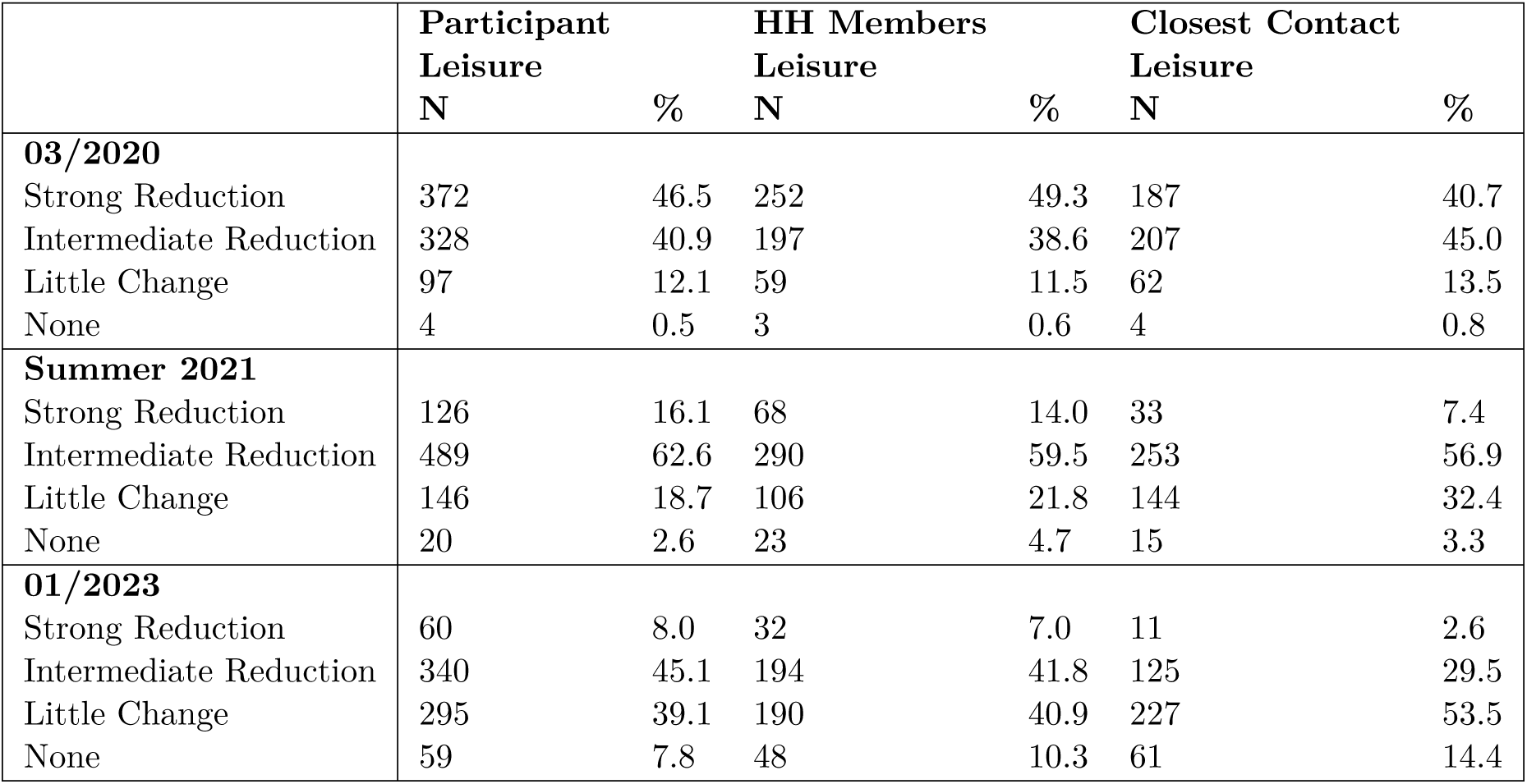
Group assignment based on the change of no. of contacts (for details see Subsection Statistical Methods). Across the three considered time points, the shares for the participants and the share for the household members are comparable. More participants than closest contacts are assigned to the “completely reduced” group.

|  | Participant Leisure |  | HH Members Leisure |  | Closest Contact Leisure |  |
| --- | --- | --- | --- | --- | --- | --- |
|  | N | % | N | % | N | % |
| **03/2020** |  |  |  |  |  |  |
| Strong Reduction | 372 | 46.5 | 252 | 49.3 | 187 | 40.7 |
| Intermediate Reduction | 328 | 40.9 | 197 | 38.6 | 207 | 45.0 |
| Little Change | 97 | 12.1 | 59 | 11.5 | 62 | 13.5 |
| None | 4 | 0.5 | 3 | 0.6 | 4 | 0.8 |
| **Summer 2021** |  |  |  |  |  |  |
| Strong Reduction | 126 | 16.1 | 68 | 14.0 | 33 | 7.4 |
| Intermediate Reduction | 489 | 62.6 | 290 | 59.5 | 253 | 56.9 |
| Little Change | 146 | 18.7 | 106 | 21.8 | 144 | 32.4 |
| None | 20 | 2.6 | 23 | 4.7 | 15 | 3.3 |
| **01/2023** |  |  |  |  |  |  |
| Strong Reduction | 60 | 8.0 | 32 | 7.0 | 11 | 2.6 |
| Intermediate Reduction | 340 | 45.1 | 194 | 41.8 | 125 | 29.5 |
| Little Change | 295 | 39.1 | 190 | 40.9 | 227 | 53.5 |
| None | 59 | 7.8 | 48 | 10.3 | 61 | 14.4 |

**Supplementary Table 8:** Output of Kolmogorov Smirnov tests comparing the ECDF of change of number of contacts between participants, their household members, and their CC. Statistic D takes the largest absolute difference between the two empirical cumulative distribution functions. Stars indicate that distributions differ at a significance level of \*\*\**p* < 0.01, \*\**p* < 0.05, \**p* < 0.1.

|  | Participant vs HH members |  | Participant vs CC |  |
| --- | --- | --- | --- | --- |
|  | D | p-value | D | p-value |
| **Work** |  |  |  |  |
| 03/2020 | 0.14 | $1.16e-05^{***}$ | 0.43 | $< 2.2e-16^{***}$ |
| Summer 2021 | 0.26 | $4.09e-16^{***}$ | 0.42 | $< 2.2e-16^{***}$ |
| 01/2023 | 0.26 | $< 2.2e-16^{***}$ | 0.43 | $< 2.2e-16^{***}$ |
| **Leisure** |  |  |  |  |
| 03/2020 | 0.06 | 0.24 | 0.42 | $< 2.2e-16^{***}$ |
| Summer 2021 | 0.12 | $4.2e-04^{***}$ | 0.41 | $< 2.2e-16^{***}$ |
| 01/2023 | 0.12 | $6.3e-04^{***}$ | 0.41 | $< 2.2e-16^{***}$ |

### Kolmogorov-Smirnov tests

#### Whole Sample

Kolmogorov-Smirnov tests were used to compare contact reductions between time points. The distributions of work and leisure contact reductions differed significantly between time points (*p* < 0.01, Supplementary Table 9).

**Supplementary Table 9:** Output of Kolmogorov Smirnov tests comparing the contact reductions between time points. Statistic D takes the largest absolute difference between the two empirical cumulative distribution functions. Stars indicate that distributions differ at a significance level of \*\*\**p* < 0.01.

|  | D | p-value |
| --- | --- | --- |
| **Work** |  |  |
| 03/2020 vs Summer 2021 | 0.21 | $< 2.2e-16^{***}$ |
| 03/2020 vs 01/2023 | 0.35 | $< 2.2e-16^{***}$ |
| Summer 2021 vs 01/2023 | 0.22 | $< 2.2e-16^{***}$ |
| **Leisure** |  |  |
| 03/2020 vs Summer 2021 | 0.32 | $< 2.2e-16^{***}$ |
| 03/2020 vs 01/2023 | 0.48 | $< 2.2e-16^{***}$ |
| Summer 2021 vs 01/2023 | 0.29 | $< 2.2e-16^{***}$ |

### Risk-Averse vs Risk-Tolerant Participants

Kolmogorov-Smirnov tests were used to compare pre-pandemic sociability between risk perception groups. In 2019, the distributions of work contacts and leisure contacts did not differ significantly between risk perception groups (*p* > 0.1, Supplementary Table 10).

The difference in distribution of work contact reductions between risk perception groups was only statistically significant in 03/2020 (*p* < 0.05, Supplementary Table 11). In the leisure context, however, the difference was statistically significant for all three subsequent time points (*p* < 0.01).

**Supplementary Table 10:** Output of Kolmogorov Smirnov tests comparing the absolute number of contacts between risk-averse and risk-tolerant participants for 2019. Statistic D takes the largest absolute difference between the two empirical cumulative distribution functions. Stars indicate that distributions differ at a significance level of \*\*\**p* < 0.01, \*\**p* < 0.05, \**p* < 0.1.

|  | D | p-value |
| --- | --- | --- |
| **Work** |  |  |
| 2019 | 0.14 | 0.45 |
| **Leisure** |  |  |
| 2019 | 0.08 | 0.94 |

**Supplementary Table 11:** Output of Kolmogorov Smirnov tests comparing the distribution of change of number of contacts between risk-averse and risk-tolerant participants. Statistic D takes the largest absolute difference between the two empirical cumulative distribution functions. Stars indicate that distributions differ at a significance level of \*\*\**p* < 0.01, \*\**p* < 0.05, \**p* < 0.1.

|  | D | p-value |
| --- | --- | --- |
| **Work** |  |  |
| 03/2020 | 0.26 | 0.01** |
| Summer 2021 | 0.19 | 0.12 |
| 01/2023 | 0.17 | 0.21 |
| **Leisure** |  |  |
| 03/2020 | 0.32 | 6.9e-04*** |
| Summer 2021 | 0.30 | 9.5e-04*** |
| 01/2023 | 0.30 | 0.002*** |

### Subgroup Analysis by Age Group

Relative to 2019, all three age groups reduced their work contacts in 03/2020, summer 2021, and 01/2023. All three age groups relaxed their mean reductions of work contacts over time: In 03/2020, 18-39 year olds reduced their work contacts on average by 83%, 40-59 year olds by 75%, and 60+ year olds by 63% (Supplementary Figure 11). In summer 2021, mean work contact reductions were relaxed to 69% (18-39 year olds), 63% (40-59 year olds), and 61% (60+ year olds). Finally, in 01/2023, they were relaxed to 46% (18-39 year olds), 39% (40-59 year olds), and 49% (60+ year olds). For all three time points, the difference in distribution between 18-39 year olds and 60+ year olds and between 40-59 year olds and 60+ year olds is statistically significant (*p* < 0.05 for some comparisons, *p* < 0.01 for others, see Supplementary Table 14). Just like for the whole sample, the Bayesian reduction model confirms the temporal development of the reductions: The share of participants who “strongly” reduced their work contacts decreased over time for all three age groups, while the share of participants who displayed “little change” in their number of work contacts increased (Supplementary Table 13). In sum, although all age groups reduced their work contacts substantially at the beginning of the COVID-19 pandemic, work contact reductions were incrementally relaxed over time, with 18-39 year olds consistently showing the highest reductions and 60+ year olds the lowest.

Considering the change of number of leisure contacts, we observe that, on average all three age groups reduced their leisure contacts in 03/2020, summer 2021, and 01/2023. For 03/2020, 18-39 year olds reported an average reduction of 78%, 40-59 year olds of 73%, and 60+ year olds of 62% (Supplementary Figure 12). From 03/2020 to summer 2021 these reductions were relaxed to 46% (18-39 year olds), 52% (40-59 year olds), and 46% (60+ year olds). In 01/2023 mean reductions had been further relaxed, to 29% (18-30 year olds), 27% (40-59 year olds), and 30% (60+ year olds). The comparability and time-dependence of mean reduction is once more confirmed by the results of the Bayesian reduction model: For all three time points, the group sizes across age groups are comparable. The difference in distribution is only statistically significant for 40-59 year olds and 60+ year olds in 03/2020 (*p* < 0.1, Supplementary Table 14). In sum, all three age groups comparably reduced their leisure contacts and incrementally relaxed these reductions over time.

**Supplementary Table 12:** Age group sizes. The majority of the participants was between 40 and 59 years old. Participants had to be of legal age (18+) to qualify for participation.

| Age category | N | Percent |
| --- | --- | --- |
| 18-39 | 155 | 18.2 |
| 40-59 | 566 | 66.6 |
| 60+ | 129 | 15.2 |
| Missing | 13 | — |

**Supplementary Table 13:** Group assignment based on the change of no. of contacts (for details see Subsection Statistical Methods). All age groups tended to reduce their work contacts more than their leisure contacts. In 03/2020, the 18-39 year olds had the largest share of participants who completely cut their work contacts, while by 01/2023 they had the smallest share of participants who completely reduced. For leisure, shares are comparable across age groups.

|  | Work<br>18-39 |  | Work<br>40-59 |  | Work<br>60+ |  | Leisure<br>18-39 |  | Leisure<br>40-59 |  | Leisure<br>60+ |  |
| --- | --- | --- | --- | --- | --- | --- | --- | --- | --- | --- | --- | --- |
|  | N | % | N | % | N | % | N | % | N | % | N | % |
| **03/2020** |  |  |  |  |  |  |  |  |  |  |  |  |
| Strong Reduction | 88 | 64.7 | 269 | 54.7 | 38 | 42.2 | 69 | 47.6 | 260 | 49.3 | 45 | 38.1 |
| Intermediate Reduction | 33 | 24.3 | 138 | 28.0 | 29 | 32.2 | 66 | 45.5 | 201 | 38.2 | 49 | 41.5 |
| Little Change | 15 | 11.0 | 82 | 16.7 | 20 | 22.2 | 10 | 6.9 | 64 | 12.1 | 22 | 18.7 |
| None | 0 | 0.0 | 3 | 0.6 | 3 | 3.4 | 0 | 0.0 | 2 | 0.4 | 2 | 1.7 |
| **Summer 2021** |  |  |  |  |  |  |  |  |  |  |  |  |
| Strong Reduction | 57 | 43.2 | 200 | 41.5 | 37 | 40.2 | 20 | 14.4 | 83 | 16.0 | 19 | 16.8 |
| Intermediate Reduction | 58 | 43.9 | 188 | 39.0 | 33 | 35.9 | 86 | 61.9 | 332 | 64.2 | 64 | 56.6 |
| Little Change | 17 | 12.9 | 88 | 18.3 | 18 | 19.6 | 28 | 20.1 | 97 | 18.8 | 20 | 17.7 |
| None | 0 | 0.0 | 6 | 1.2 | 4 | 4.3 | 5 | 3.6 | 5 | 1.0 | 10 | 8.9 |
| **01/2023** |  |  |  |  |  |  |  |  |  |  |  |  |
| Strong Reduction | 31 | 23.3 | 94 | 19.7 | 35 | 38.5 | 15 | 11.0 | 36 | 7.1 | 11 | 10.8 |
| Intermediate Reduction | 62 | 46.6 | 193 | 40.4 | 24 | 26.4 | 61 | 44.8 | 227 | 45.0 | 46 | 45.1 |
| Little Change | 34 | 25.6 | 166 | 34.7 | 26 | 28.6 | 44 | 32.4 | 208 | 41.3 | 35 | 34.3 |
| None | 6 | 4.5 | 25 | 5.2 | 6 | 6.5 | 16 | 11.8 | 33 | 6.6 | 10 | 9.8 |

The lack of difference in reduction of leisure contacts is reflected by the lack of difference in reported number of infections: Of the 18-39 year olds 34% (95% CI [26%, 41%]) reported zero infections vs 32% (95% CI [28%, 36%]) of the 40-59 year olds and 43% (95% CI [34%, 52%]) of the 60+ year olds (Supplementary Figure 13 A). Consequently, the share of participants who reported one infection was smallest for the 60+ year olds (53%, 95% CI[44%, 62%]), followed by the 18-39 year olds (56%, 95% CI [48%, 63%]), and the 40-59 year olds (57%, 95% CI [52%, 60%]). Two infections were reported by 9% (95% CI [5%, 13%]) of 18-39 year olds, by 10% (95% CI [8%, 13%]) of 40-59 year olds, and by 5% (95% CI [1%, 8%]) of 60+ year olds. The difference in distribution of number of reported infections is statistically insignificant (*p* > 0.1). The ECDFs of the timing of the first infection of the three age groups also do not display any significant difference (*p* > 0.1, Supplementary Figure 13 B). Overall, the two younger age groups reduced their work contacts more strongly than the 60+ year olds, but this difference in reduction can neither be found for reduction of leisure contacts, nor for the number of infections or the timing of first infection.

**Supplementary Figure 11:**
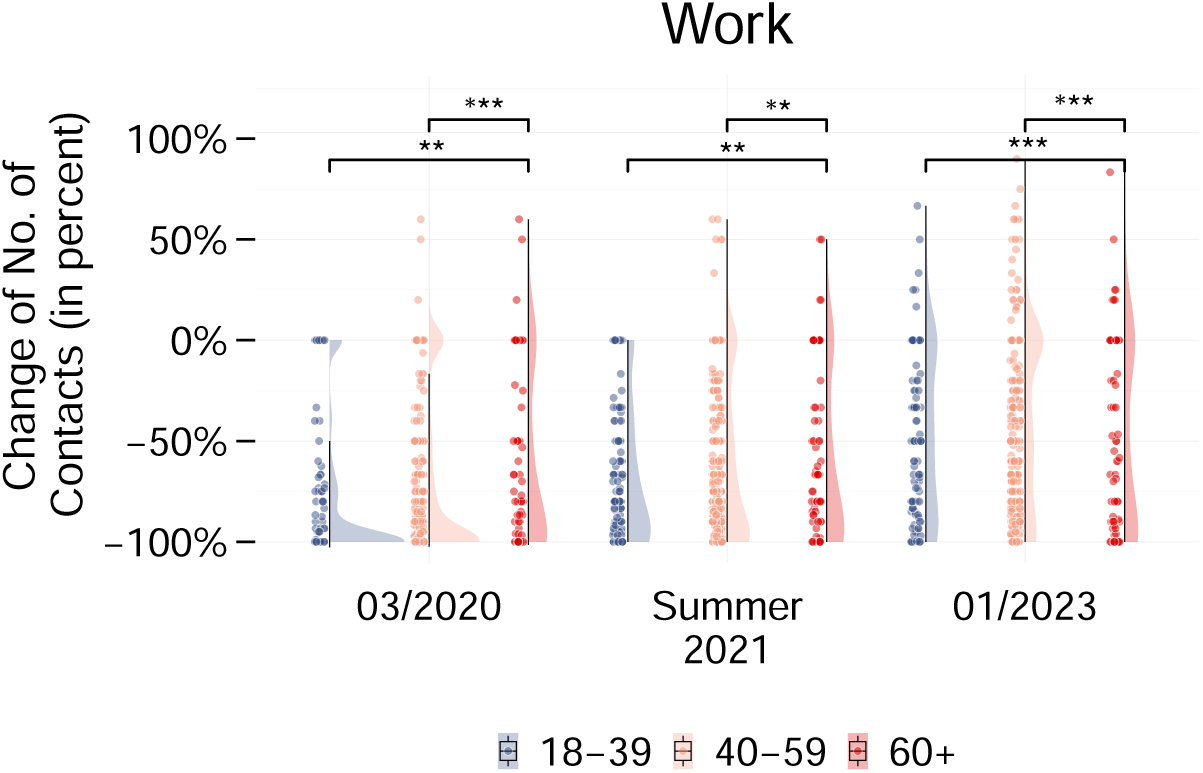
Change of number of work contacts (in percent, relative to 2019) for different age groups. Relative number of work contacts. In March 2020, the share of 18-39 year old participants who “strongly” reduced their work contacts was larger than the corresponding share of 40-59 year old and 60+ year old participants.

**Supplementary Figure 12:**
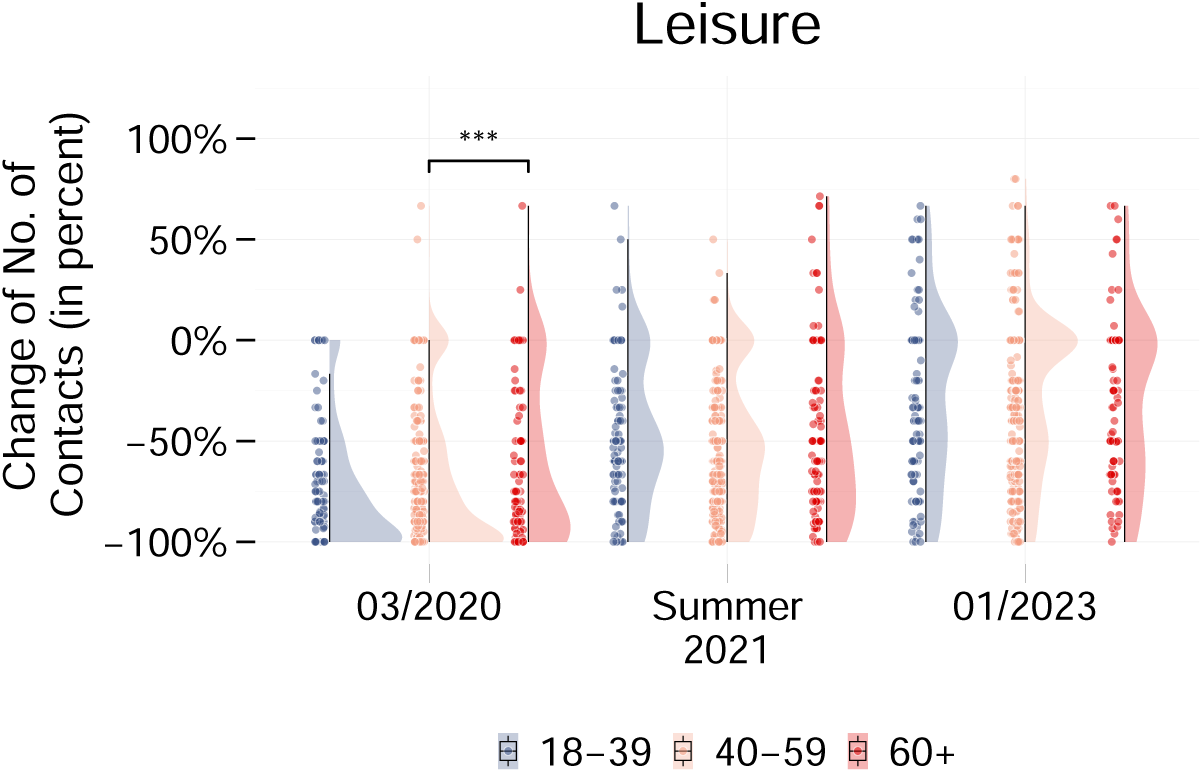
Change of number of leisure contacts (in percent, relative to 2019) for different age groups. Relative number of leisure contacts. The share of 60+ year olds who “strongly” reduced their leisure contacts is smaller than the corresponding share of 18-39 and 40-59 year olds in 03/2020. For the two later time points, reductions are comparable across age groups.

**Supplementary Figure 13:**
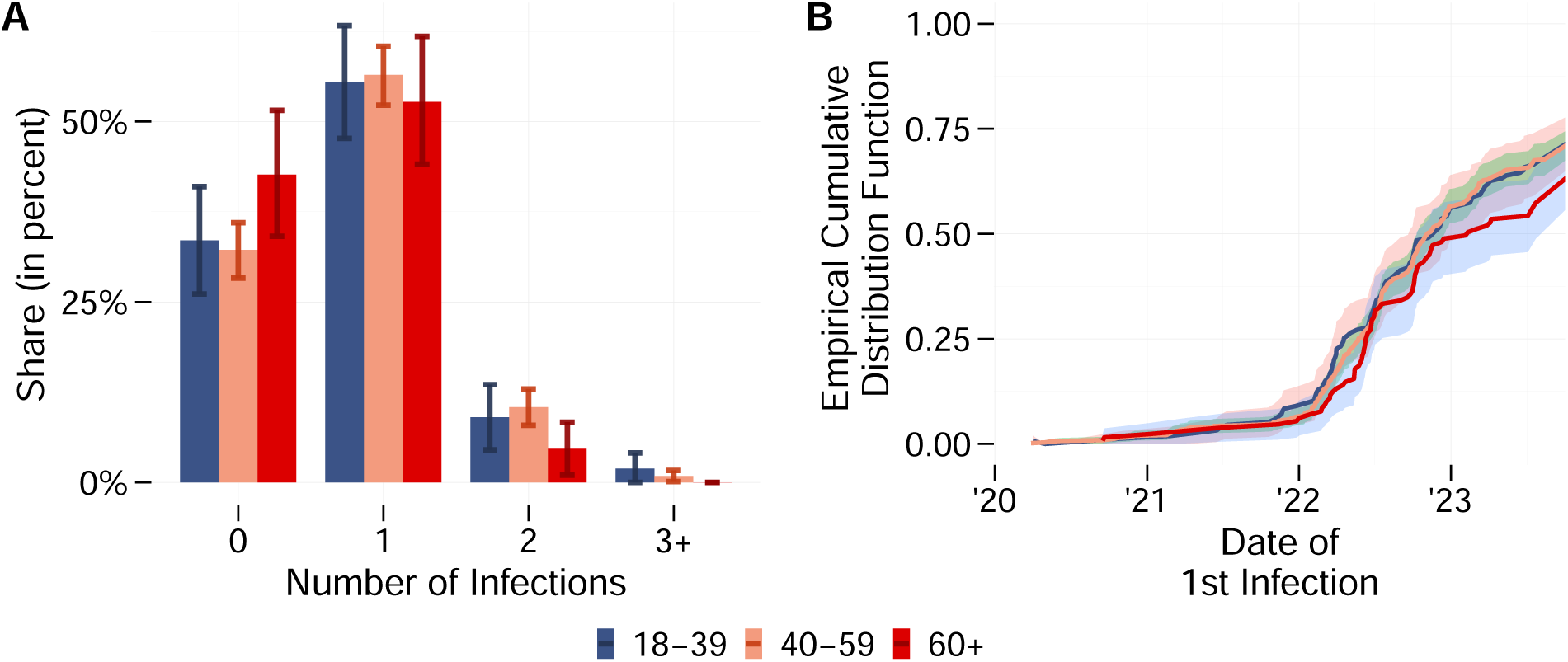
**A.** Share of respondents who reported 0/1/2/3+ infections, differentiated by age groups. Error bars represent 95% confidence intervals (see Subsection Statistical Methods for computation and motivation). The share of participants who reported no infections, is largest for 60+ year olds. **B.** Timing of participant’s (first) infection(s); differentiated by age group. Ribbons represent 95% confidence intervals (see Subsection Statistical Methods for computation and motivation).The ECDF of 60+ year olds is from mid-2021 onwards constantly below the ECDF of the other two age groups.

**Supplementary Table 14:** Output of Kolmogorov Smirnov tests comparing the ECDF of change of number of contacts between age groups. Statistic D takes the largest absolute difference between the two empirical cumulative distribution functions. Stars indicate that distributions differ at a significance level of \*\*\**p* < 0.01, \*\**p* < 0.05, \**p* < 0.1.

|  | 18-39 vs 40-59 |  | 18-39 vs 60+ |  | 40-59 vs 60+ |  |
| --- | --- | --- | --- | --- | --- | --- |
|  | D | p-value | D | p-value | D | p-value |
| **Work** |  |  |  |  |  |  |
| 03/2020 | 0.11 | 0.11 | 0.19 | 0.01** | 0.18 | 0.003*** |
| Summer 2021 | 0.06 | 0.67 | 0.18 | 0.02** | 0.14 | 0.02** |
| 01/2023 | 0.09 | 0.35 | 0.25 | 0.0003*** | 0.25 | 4.75e-06*** |
| **Leisure** |  |  |  |  |  |  |
| 03/2020 | 0.05 | 0.94 | 0.15 | 0.11 | 0.13 | 0.08* |
| Summer 2021 | 0.11 | 0.11 | 0.15 | 0.12 | 0.07 | 0.78 |
| 01/2023 | 0.07 | 0.63 | 0.04 | 1.0 | 0.06 | 0.88 |

### Subgroup Analysis by Gender

Relative to 2019, both genders reduced their work contacts in 03/2020, in summer 2021, and in 01/2023. Both genders relaxed their work reductions over time: Relative to 2019, female participants reported an average reduction of work contacts of 75% (male participants: 74%) in 03/2020, of 61% (male participants: 67%) in summer 2021, and of 39% (male participants: 44%) in 01/2023 (Supplementary Figure 14 A). This negligibly small difference between genders is reflected by the group assignments of the Bayesian reduction model: With the exception of summer 2021, when a slightly smaller share of female than male participants “strongly” reduced their work contacts (41% vs 44%), hardly any difference between the genders can be noted (Supplementary Table 16). Kolmogorov-Smirnov-tests classify the difference in distribution of change of number of work contacts as statistically insignificant for all time points (*p* > 0.1, Supplementary Table 17).

**Supplementary Table 15:** Gender distribution of the survey participants. As people who reported their gender as “diverse” make up less than 1% of the sample size and only include 7 people, they were excluded from the subanalysis.

| Gender | N | Percent |
| --- | --- | --- |
| Female | 464 | 54.2 |
| Male | 385 | 45.0 |
| Diverse | 7 | 0.8 |
| Missing | 10 | — |

**Supplementary Figure 14:**
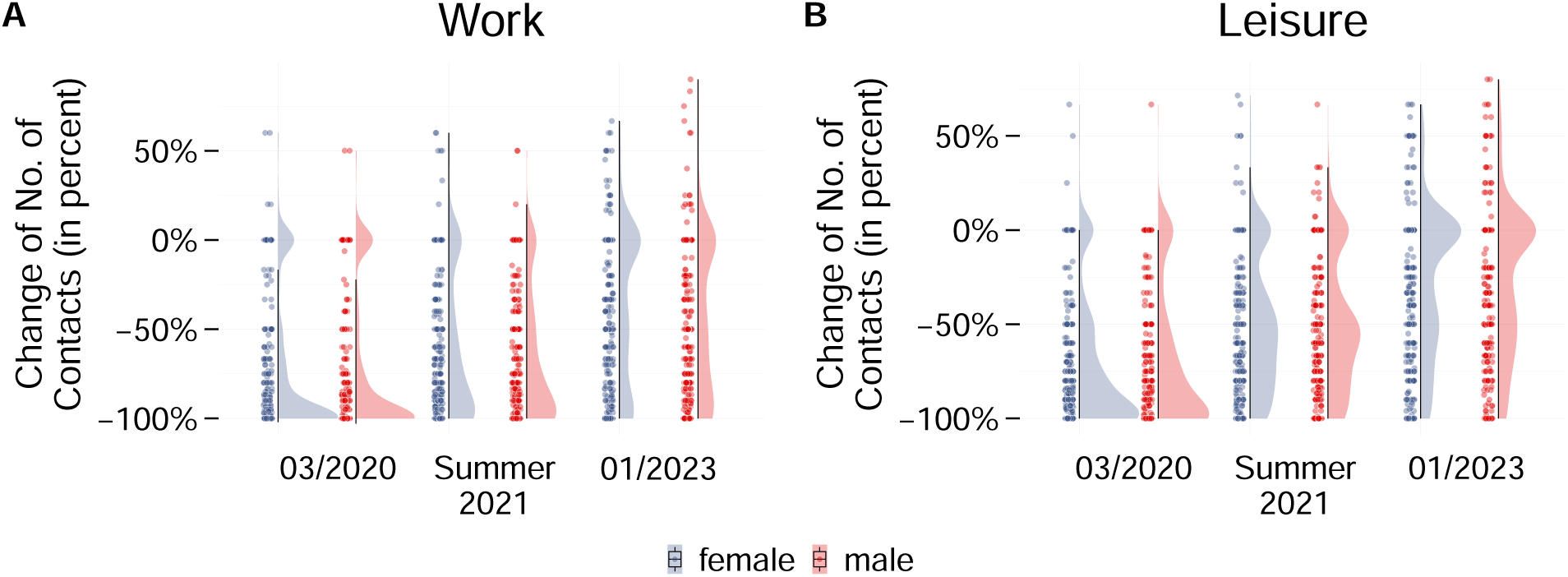
Change of number of contacts (in percent, relative to 2019) for different genders. As a percentage increase from zero cannot be computed, individuals who reported zero work/leisure contacts in 2019, were excluded from the corresponding context. **A.** Relative number of work contacts. Both gender reported comparable reductions of work contacts. **B.** Relative number of leisure contacts. Both gender reported comparable reductions of leisure contacts.

Analogously to their work contacts, both genders reduced their leisure contacts at all three time points. Female participants reported an average reduction of 73% (male participants: 71%) in 03/2020, of 50% (male participants: 50%) in summer 2021, and of 28% (male participants: 26%) in 01/2023 (Supplementary Figure 14 B). The results of the Bayesian reduction model are comparable (Supplementary Table 16) and according to the Kolmogorov-Smirnov-Test the difference in distribution did not reach statistical significance at any time point (*p* > 0.1, Supplementary Table 17).

Comparing the number of reported infections reveals that 36% of female participants (95% CI: [31%,40%]) vs 31% of male participants (95% CI: [27%,26%]) reported never having been infected (Supplementary Figure 15 A). As a consequence, the share of female participants who reported one infection is slightly smaller. Female participants: 54% (95% CI: [49%,58%]), male participants: 58% (95% CI: [54%,63%]). The difference in distribution, however, is not statistically significant (p*>* 0.1, Supplementary Table 17). Analogously, the ECDFs of the timing of the first infection for female and male participants coincide (Supplementary Figure 15 B). Again, the difference in distribution is not statistically significant (p > 0.1).

**Supplementary Table 16:** Group assignment based on the change of no. of contacts (for details see Subsection Statistical Methods). Both genders reduced their work contacts more strongly than their leisure contacts. With the exception of summer 2021, for which more male than female participants were assigned to the “strong reduction” group, hardly any differences between the genders can be noted.

|  | **Work**<br>**Female** |  | **Work**<br>**Male** |  | **Leisure**<br>**Female** |  | **Leisure**<br>**Male** |  |
| --- | --- | --- | --- | --- | --- | --- | --- | --- |
|  | N | % | N | % | N | % | N | % |
| **03/2020** |  |  |  |  |  |  |  |  |
| Strong Reduction | 212 | 54.4 | 180 | 55.6 | 205 | 47.2 | 162 | 46.0 |
| Intermediate Reduction | 113 | 29.0 | 87 | 26.9 | 180 | 41.5 | 141 | 40.1 |
| Little Change | 61 | 15.6 | 55 | 17.0 | 46 | 10.6 | 48 | 13.6 |
| None | 4 | 1.0 | 2 | 0.5 | 3 | 0.7 | 1 | 0.3 |
| **Summer 2021** |  |  |  |  |  |  |  |  |
| Strong Reduction | 155 | 40.7 | 142 | 44.2 | 70 | 16.7 | 52 | 15.1 |
| Intermediate Reduction | 142 | 37.3 | 131 | 40.8 | 261 | 62.1 | 218 | 63.2 |
| Little Change | 77 | 20.2 | 45 | 14.0 | 80 | 19.0 | 65 | 18.8 |
| None | 7 | 1.8 | 3 | 1.0 | 9 | 2.2 | 10 | 2.9 |
| **01/2023** |  |  |  |  |  |  |  |  |
| Strong Reduction | 83 | 22.1 | 73 | 22.6 | 34 | 8.4 | 25 | 7.5 |
| Intermediate Reduction | 140 | 37.2 | 139 | 43.0 | 177 | 43.8 | 152 | 45.5 |
| Little Change | 131 | 34.8 | 96 | 29.7 | 163 | 40.3 | 129 | 38.6 |
| None | 22 | 5.9 | 15 | 4.7 | 30 | 7.5 | 28 | 8.4 |

**Supplementary Figure 15:**
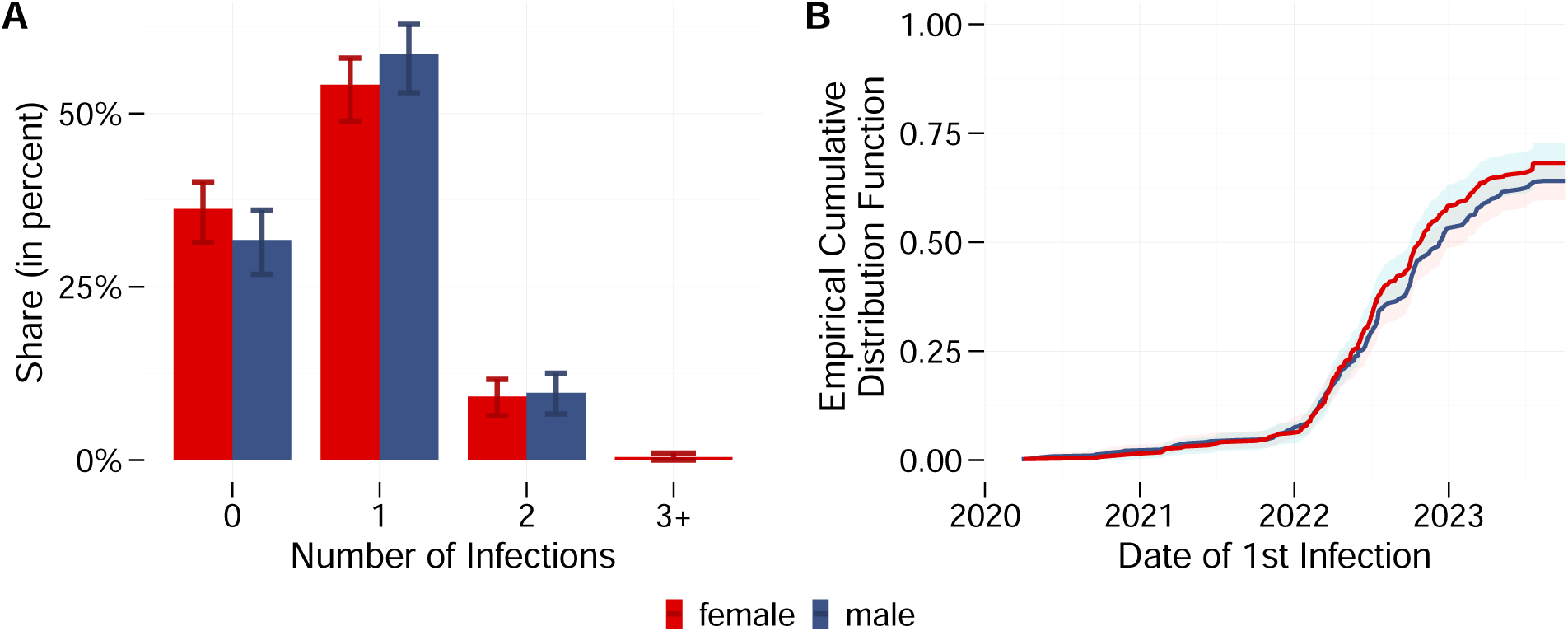
**A.** Share of respondents who reported 0/1/2/3+ infections, differentiated by gender. Error bars represent 95% confidence intervals (see Subsection Statistical Methods for computation and motivation). Female participants reported less infections than male participants. **B.** Number and timing of respondent’s (first) infection(s); differentiated by gender. Ribbons represent 95% confidence intervals (see Subsection Statistical Methods for computation and motivation).

**Supplementary Table 17:**
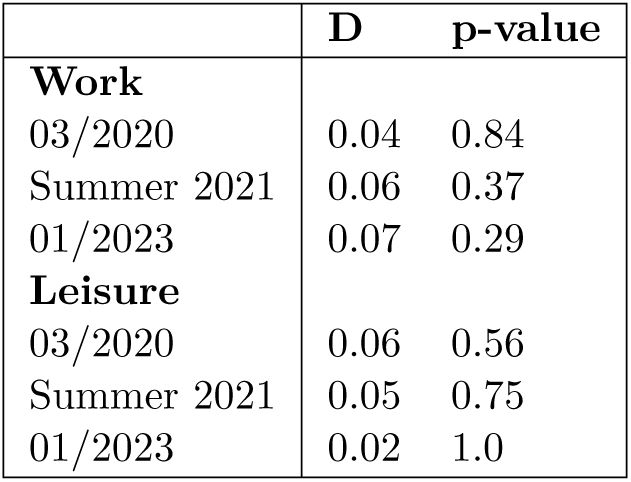
Output of Kolmogorov Smirnov tests comparing the ECDF of change of number of contacts between female and male participants. Statistic D takes the largest absolute difference between the two empirical cumulative distribution functions. For no time point and neither context does the test deem the difference in distribution statistically significant.

|  | **D** | **p-value** |
| --- | --- | --- |
| **Work** |  |  |
| 03/2020 | 0.04 | 0.84 |
| Summer 2021 | 0.06 | 0.37 |
| 01/2023 | 0.07 | 0.29 |
| **Leisure** |  |  |
| 03/2020 | 0.06 | 0.56 |
| Summer 2021 | 0.05 | 0.75 |
| 01/2023 | 0.02 | 1.0 |

### Subgroup Analysis by Comorbidities

The survey gave participants the possibility to report the following comorbidities:

- high blood pressure (hypertension),
- diabetes (diabetes mellitus),
- cardiovascular disease (e.g. coronary heart disease, any condition following a heart attack, heart failure, cardiac arrhythmia, any condition following a stroke),
- chronic lung disease (e.g. asthma, chronic bronchitis, chronic obstructive pulmonary disease (COPD), emphysema),
- current immunodeficiency (e.g. due to an illness, organ transplant, chemotherapy or currently taking other medication such as cortisone),
- cancer for which you are currently being treated or have been treated in the last year,
- post-COVID-19 condition.

**Supplementary Table 18:** Number and shares of participants who reported no/some comorbidity.

| Subgroup | N | Percent |
| --- | --- | --- |
| No Comorbidity | 593 | 68.8 |
| Some Comorbidity | 269 | 31.2 |
| Missing | 5 | — |

As individuals with comorbidities are at a higher risk of developing severe COVID-19, they may reduce their social contacts stronger than the general population [DMFdPCV22]. In our sample, however, participants without comorbidity and participants with some comorbidity reduced their work and leisure contacts comparably. In 03/2020, participants without comorbidities reduced their work contacts on average by 76% (some comorbidity: 71%) and their leisure contacts on average by 73% (some comorbidity: 72%). Both groups relaxed their reduction over time, leading to an average work reduction of 65% for participants without comorbidity and 61% for participants with some comorbidity in summer 2021. In the leisure context, the difference in average reduction is even slimmer in summer 2021 with 51% (no comorbidities) vs 50% (some comorbidity). Relaxations of reductions from summer 2021 to 01/2023 are similar for both groups (Supplementary Figure 16). For 03/2020 and summer 2021, the lack of difference is confirmed by the Bayesian reduction model (Supplementary Table 19). In 01/2023, however, a larger share of participants without comorbidity showed strong reduction (*p* < 0.05, Supplementary Table 20). Overall, participants with and without comorbidity reduced contacts similarly in 03/2020 and summer 2021, while in 01/2023, participants without comorbidity reduced more strongly.

This lack of difference in contact reductions propagates to a lack of difference in number of reported infections: 35% (95% CI: [32%, 38%]) of participants without comorbidity reported no infections vs 32% (95% CI: [28%, 36&]) of participants with comorbidity. This small difference then leads to a small difference in share of participants who reported one infection (no comorbidity: 55%, 95% CI: [50%, 59%], some comorbidity: 58%, 95% CI: [51%, 65%]). The share of participants who reported two infections is comparable (no comorbidities: 9%, 95% CI: [9%, 10%], some comorbidity: 9%, 95% CI: [8%, 10 %]). Less than 1% of either group reported having been infected at least three times (Supplementary Figure 17 A). The difference in distribution is not statistically significant (*p* > 0.1). Finally,the ECDFs of the timing of the first infection for participants with/without comorbiditiy are indistinguishable (Supplementary Figure 17 B, *p* > 0.1).

**Supplementary Figure 16:**
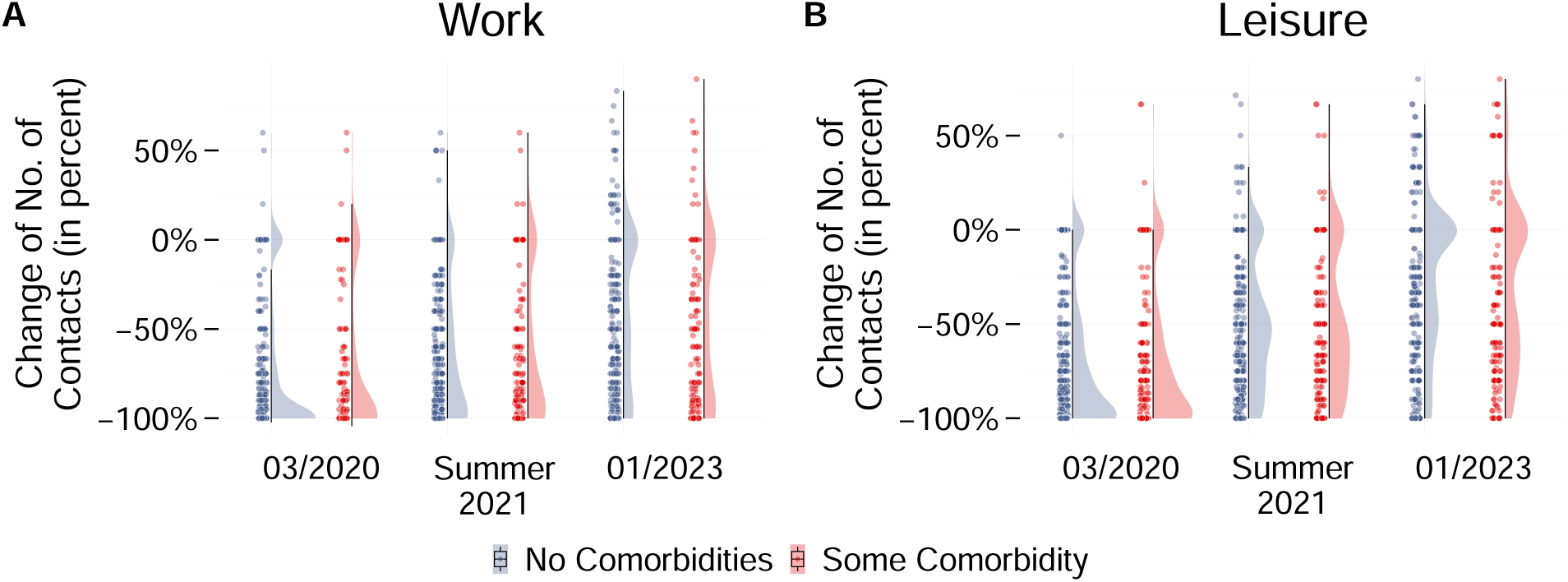
Violin plots of change of number of contacts (relative to 2019) by with/without comorbidity. The difference in mean is not significant for the three points in time and for both work and leisure contacts.

**Supplementary Table 19:** Group assignment based on the change of no. of contacts (for details see Subsection Statistical Methods). Both groups tended to reduce their work contacts more than their leisure contacts.

|  | **Work**<br>**No Com.** |  | **Work**<br>**Some Com.** |  | **Leisure**<br>**No Com.** |  | **Leisure**<br>**Some Com.** |  |
| --- | --- | --- | --- | --- | --- | --- | --- | --- |
|  | N | % | N | % | N | % | N | % |
| **03/2020** |  |  |  |  |  |  |  |  |
| Strong Reduction | 282 | 55.6 | 113 | 52.3 | 254 | 46.4 | 112 | 45.0 |
| Intermediate Reduction | 145 | 28.6 | 59 | 27.3 | 228 | 41.6 | 102 | 41.0 |
| Little Change | 77 | 15.2 | 41 | 19.0 | 65 | 11.9 | 32 | 12.9 |
| None | 3 | 0.6 | 3 | 1.4 | 1 | 0.1 | 3 | 1.1 |
| **Summer 2021** |  |  |  |  |  |  |  |  |
| Strong Reduction | 206 | 41.7 | 94 | 43.7 | 82 | 15.4 | 42 | 17.4 |
| Intermediate Reduction | 212 | 42.7 | 66 | 30.7 | 345 | 64.6 | 141 | 58.3 |
| Little Change | 72 | 14.5 | 51 | 23.7 | 94 | 17.6 | 52 | 21.5 |
| None | 6 | 1.3 | 4 | 1.9 | 13 | 2.4 | 7 | 2.8 |
| **01/2023** |  |  |  |  |  |  |  |  |
| Strong Reduction | 50 | 36.0 | 60 | 28.4 | 44 | 8.5 | 16 | 6.8 |
| Intermediate Reduction | 40 | 28.8 | 73 | 34.6 | 223 | 43.3 | 113 | 48.3 |
| Little Change | 40 | 28.8 | 69 | 32.7 | 209 | 40.6 | 85 | 36.3 |
| None | 9 | 6.4 | 9 | 4.3 | 39 | 7.6 | 20 | 8.6 |

**Supplementary Figure 17:**
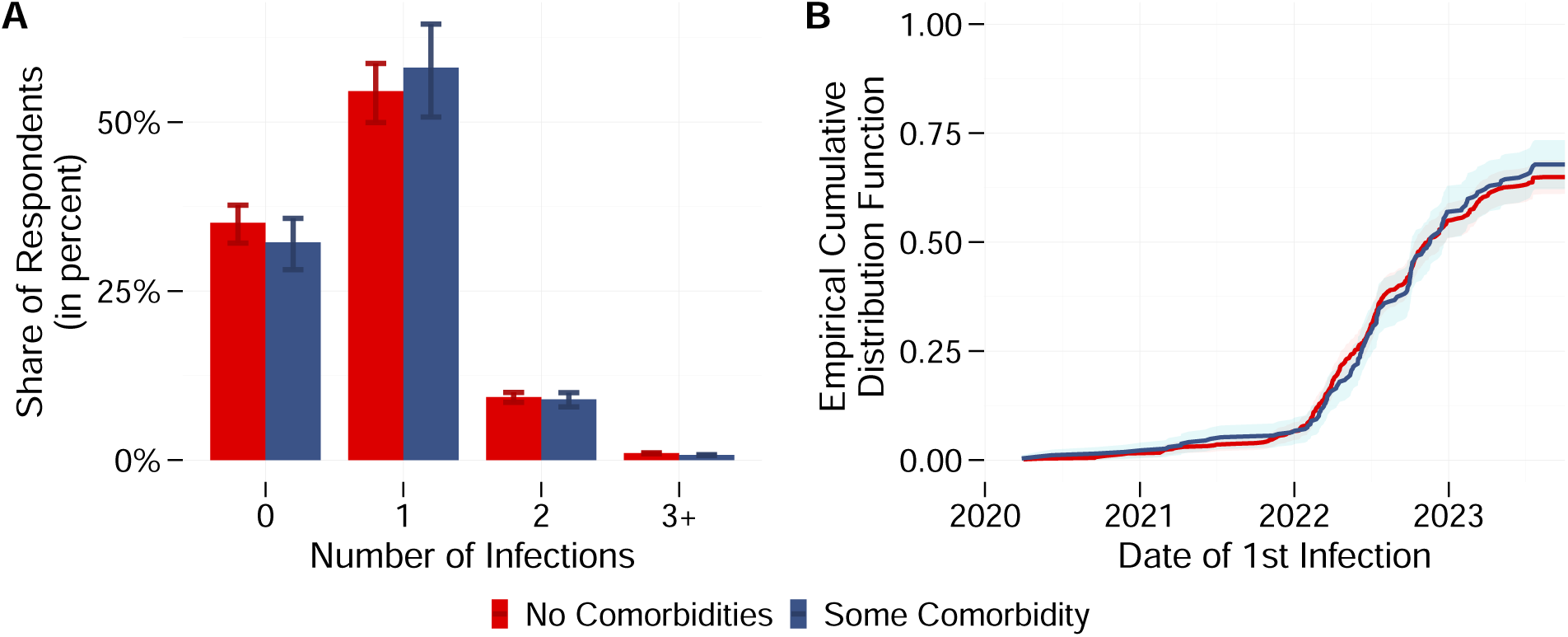
**A.**Share of respondents who reported 0/1/2/3+ infections, differentiated by with/withour comorbidity. Error bars represent 95% confidence intervals (see Subsection Statistical Methods for computation and motivation). **B.** Number and timing of participant’s (first) infection(s); differentiated by whether they reported a COVID-19 relevant comorbidity. Ribbons represent 95% confidence intervals (see Subsection Statistical Methods for computation and motivation).

**Supplementary Table 20:** Output of Kolmogorov Smirnov tests comparing the ECDF of change of number of contacts between participants without/with some comorbiditiy. Statistic D takes the largest absolute difference between the two empirical cumulative distribution functions. Stars indicate that distributions differ at a significance level of \*\*\**p* < 0.01, \*\**p* < 0.05, \**p* < 0.1.

|  | D | p-value |
| --- | --- | --- |
| **Work** |  |  |
| 03/2020 | 0.05 | 0.75 |
| Summer 2021 | 0.08 | 0.18 |
| 01/2023 | 0.11 | 0.02** |
| **Leisure** |  |  |
| 03/2020 | 0.02 | 1.00 |
| Summer 2021 | 0.06 | 0.60 |
| 01/2023 | 0.10 | 0.07* |

